# Associations between loneliness and outcomes of Common Mental Disorders (CMDs): A systematic review of longitudinal studies

**DOI:** 10.1101/2025.08.06.25333144

**Authors:** Theodora Stefanidou, Gamze Evlat, Hannah Gray, Kinga Lagan, Jasmine Harju-Seppänen, Millie Osullivan, Shivangi Talwar, Gemma Lewis, Joshua E J Buckman, Alexandra Pitman, Sonia Johnson, Brynmor Lloyd-Evans

## Abstract

**Background:** Loneliness has been increasingly associated with poor mental health outcomes, yet its prognostic role in people with Common Mental Disorders (CMDs) remains unclear. This systematic review aimed to examine whether loneliness is longitudinally associated with mental health outcomes in individuals with CMDs, regardless of treatment status.

**Methods:** We searched PsycINFO, MEDLINE, Embase and CINAHL from inception to May 2024 for longitudinal studies examining associations between baseline loneliness and CMD outcomes at follow-up. Eligible studies included adults aged 16+ with CMDs. Study quality was assessed using the Newcastle-Ottawa Scale, and certainty of evidence was evaluated using GRADE. Due to heterogeneity in population and outcomes, a narrative synthesis was conducted. The protocol was registered with PROSPERO (registration number: CRD42023410401).

**Results:** Seventeen studies met the inclusion criteria. Most included studies investigated depression-related outcomes (n=13), with fewer addressing suicidal ideation (n=5), anxiety (n=2), and mixed CMD outcomes (n=1). Several studies contributed data to more than one outcome category. We found that high baseline loneliness was consistently associated with poorer depression-related outcomes at follow-up in people with CMDs. This association was observed across clinical, community, and treatment settings. In contrast, evidence for suicidal ideation and anxiety outcomes was limited and mixed, with inconsistent associations and lower study quality. Only two studies examined treatment outcomes, with mixed findings on whether loneliness influenced intervention response. Overall, the GRADE certainty of evidence was high for depression, low for suicidal ideation, and very low for anxiety and mixed CMD outcomes.

**Conclusions:** Loneliness appears to be a consistent prognostic factor for poor depression outcomes in people with CMDs, highlighting its potential relevance for assessment and intervention. However, its role in anxiety, suicidality, and treatment response remains unclear. Further high-quality longitudinal research is needed to clarify whether—and under what conditions—loneliness affects broader CMD outcomes and treatment effectiveness.

## Background

### Conceptualisations of Loneliness

Loneliness is a distressing emotional state stemming from the subjectively experienced gap between one’s actual and desired social relations [1]. It is related to perceived social isolation and is associated with the quality of social relationships [2]. While often associated with social isolation, loneliness extends beyond physical solitude and should be distinguished from quantitative features of social support, such as objective social isolation and social network size [3].

The sociologist Weiss [4] suggested two primary dimensions of loneliness: social and emotional. According to this typology, social loneliness results from difficulties with social integration, and emotional loneliness is derived from the absence of a close intimate relationship. Expanding on Weiss’s framework, Cacioppo et al. [5] conceptualised loneliness as comprising three dimensions: intimate, relational, and collective loneliness. Intimate loneliness refers to the absence of a close emotional bond (partner, best friend), relational loneliness refers to the lack of quality friendships or infrequent contact with friends/family, and collective loneliness entails the lack of connection to wider social identities or communities (e.g., clubs, religious, or cultural groups). These models explain loneliness as a multifaceted phenomenon with diverse manifestations across contexts and individuals.

Psychometrically robust self-report measures of loneliness have been developed and used extensively in research. The UCLA Loneliness Scale, developed by Russell et al. [6], is one of the most widely used measures, assessing subjective feelings of loneliness and social isolation. Additionally, the De Jong Gierveld Loneliness Scale offers an assessment of loneliness which encompasses both emotional and social dimensions [7].

### Loneliness and Common Mental Disorders

Loneliness is increasingly recognised as a critical public health issue with evidence of adverse effects on physical and mental health [2, 8, 9]. Chronic loneliness is associated with increased risk of various health problems, including cardiovascular disease, hypertension, weakened immune function, and elevated mortality rates [10]. Furthermore, loneliness is associated with negative mental health outcomes, including increased symptoms of Common Mental Disorders (CMDs) [5, 11] suicidal thoughts, and suicide attempt [12]. According to National Institute for Health and Care Excellence (NICE) clinical guidelines, CMDs include depression and anxiety disorders such as generalised anxiety disorder (GAD), panic disorder, mixed depression and anxiety, obsessive-compulsive disorder (OCD), post-traumatic stress disorder (PTSD), and phobias about specific things or situations [13].

Depression is a leading cause of disease burden globally [14, 15]. The bidirectional relationship between loneliness and depression is well established, with longitudinal studies finding that loneliness serves as both a risk factor for the onset of depressive symptoms and as a consequence of experiencing them [16, 17]. Similarly, anxiety disorders have been linked to experiences of loneliness [18]. A meta-analysis identified that social isolation— including loneliness—is closely associated with social anxiety disorder, although causality remained unclear due to limited longitudinal evidence [19]. Longitudinal findings from a general population sample suggest that while loneliness predicts later symptoms of depression, social anxiety, and paranoia, only social anxiety prospectively predicts subsequent loneliness, highlighting its key role in the maintenance of loneliness [20].

The relationship between loneliness and mental health extends beyond clinical outcomes, encompassing broader implications for individuals’ psychological well-being and quality of life [11]. Chronic loneliness has been associated with heightened levels of psychological distress, decreased self-esteem, and impaired interpersonal functioning, exacerbating individuals’ vulnerability to mental health disorders [21]. Loneliness has also been associated with maladaptive coping strategies, such as substance abuse and disordered eating behaviours [22].

Despite these findings, there is limited evidence on whether loneliness predicts poorer outcomes in clinical populations with CMDs. One prior systematic review examined the role of loneliness and perceived social support in relation to mental health outcomes [23], but it included only two studies that directly assessed loneliness and just five of the 34 eligible studies involved long-term follow-up. Furthermore, it remains unclear whether and how loneliness may affect the outcomes of psychological or other treatments and if people who are lonely at baseline experience poorer outcomes from mental health interventions.

### Aims

Thus, this study aimed to address this important gap in knowledge by systematically reviewing the evidence on whether loneliness is associated with subsequent poor outcomes among people with CMDs. In particular, we focused on loneliness as the sole exposure of interest, and adults of all ages with CMDs as the specific population. Our primary review question was: Is loneliness longitudinally associated with mental health outcomes among adults with CMDs?

As a secondary aim, we investigated whether being lonely is associated with poorer treatment outcomes; an important question with direct clinical relevance, especially if loneliness is not directly addressed in treatment.

The review follows guidance from the Centre for Reviews and Dissemination on undertaking reviews in health care and the Preferred Reporting Items for Systematic Reviews and Meta-analyses (PRISMA statement) [24, 25]. A PRISMA checklist for this review is provided in Appendix 1.

## Methods

### Protocol and registration

The review protocol was registered at PROSPERO international prospective register of systematic reviews at the Centre for Reviews and Dissemination, University of York (CRD42023410401) [26].

### Eligibility Criteria

a. <u>Publication types:</u> Peer-reviewed published papers, grey literature and preprints were eligible.
b. <u>Study types:</u> All longitudinal studies, including both observational and intervention studies which report a longitudinal association between loneliness at T1 and subsequent mental health outcomes at T2 were included in the review. All other types of studies such as cross-sectional studies, qualitative studies, systematic reviews, and meta-analyses were excluded. However, we considered any eligible studies that are included within relevant reviews or meta-analyses.
c. <u>Participants:</u> Studies were included if their samples consisted of adults aged 16+ years with CMDs. For this review we followed the NICE clinical guidance [13] that defines CMDs as depression and anxiety disorders such as GAD, panic disorder, mixed depression and anxiety, OCD and PTSD as well as phobias about specific things or situations. Clinical populations were included however diagnosis was made, including clinical diagnoses, structured assessment tools, self-reported assessment tools or use of relevant clinical service. Within general population samples, at least 50% of participants must have met the CMD symptom threshold, as determined by clinical cut-offs specific to each screening tool or measure. If a study reported findings separately for those meeting CMD criteria, it was included, even if the total sample did not reach 50% with CMD. We excluded studies concerning samples of children under 16 years of age, people with drug and alcohol disorders only, people with a primary diagnosis of severe mental illness (SMI) (psychosis, bipolar disorder, depression with psychotic features, personality disorders), or CMDs secondary to or following the onset of neurological or neurodegenerative disorders/diseases (e.g., traumatic brain injuries, stroke, dementias, Parkinson’s disease), or populations with organic brain disorders.
d. <u>Exposure variable(s):</u> loneliness at baseline (one time point) measured by any measure including validated quantitative scales (e.g., the De Jong Gierveld Loneliness Scale or the UCLA Loneliness Scale) and single-item measures of loneliness. Studies evaluating social isolation and related concepts but not loneliness were excluded.
e. <u>Comparator(s):</u> Potential comparators included participants reporting either no loneliness or lower levels of loneliness.
f. <u>Outcomes:</u> Studies were eligible if they assessed clinical or functional (e.g., social functioning) outcomes at follow-up. The main outcome of the review was CMD symptom severity, including measures of depression, anxiety, OCD, PTSD, and overall mood. We also included suicidality. Eligible studies were required to assess CMD symptom severity at follow-up using validated instruments. In addition to CMD symptoms, secondary outcomes included well-being, global mental health, relapse following remission, admission to hospital or crisis services, quality of life, social functioning, and personal recovery. To be eligible, studies must have assessed these outcomes at follow-up using validated instruments. Relapse must have been measured through a recorded DSM or ICD diagnosis during outpatient or inpatient contact, or other reliable and validated tools. Hospital or crisis service admissions must have included voluntary or compulsory psychiatric hospitalizations, and/or use of crisis services. Overall functioning, well-being, quality of life, and personal recovery must have been assessed using validated instruments appropriate to each construct.

### Search strategy

We searched for relevant studies through four electronic databases: PsycINFO, MEDLINE, Embase and CINAHL from inception to March 2023, combining text words and subject headings (MeSH terms) for loneliness, CMDs and study design (see Appendix 2). The search terms were adapted from key reviews: Wang et al. [23] and Surkalim et al. [27]. We consulted a specialist systematic reviewer in the CORE Unit (Centre for Outcomes and Research and Effectiveness) in the Department of Psychology and Language Sciences at University College London to ensure that our search strategy was comprehensive. The search was limited to humans, and no language or publication date restrictions were imposed.

The search was updated across all databases in May 2024. We reran the original search strategy and applied an entry date filter, which retrieves articles added to the database since our last search, regardless of their publication date. Unlike a publication date filter that only captures newly published articles, the entry date filter ensures that all recently indexed articles are included in the results. We searched for additional studies and grey literature (non-peer reviewed papers) through Google Scholar by searching the first 10 pages of search results on similar terms to those used in the database searches. We also searched relevant pre-print servers (medRxiv, PsyArXiv) for eligible unpublished studies.

Backward citation tracking was conducted by reviewing the reference lists of included studies to identify relevant papers. We reviewed the citations and made note of any records that were relevant to our review. We cross checked Rayyan (a web-based systematic review management platform) to make sure the citations have not been screened already and then screened the remaining records. This process helped ensure comprehensive coverage of related literature in the review.

### Screening

Two members of the review team (TS, GE) piloted the selection strategy. All titles and abstracts were screened, with a random 26% of the records independently screened by two reviewers (TS, GE, MO, MN, JHS, ST). All full texts were independently screened by two reviewers (ST, KL, JHS, GE, HG). Screening guides are provided in Appendix 3. Any disagreements were resolved by consensus, and when necessary, by involving a third reviewer (BLE or SJ). Records were deduplicated in Endnote and then exported to Rayyan where title and abstract as well as full-text screening were carried out. Non-English records were screened using translation tools (e.g., Google Translate) and, where appropriate, native speakers assisted with confirming eligibility and data extraction.

### Data extraction

A data extraction form was developed by TS for study characteristics and outcome data. The extraction form included data from the included studies such as, the first author, year of publication, country, study design, participants, setting, outcomes, method of analysis and results. The extraction form was piloted on 10% of included studies by two reviewers (TS, HG) and revised accordingly. Data was extracted by one reviewer (TS, HG, KL).

### Quality and certainty of evidence

The quality of included studies was assessed using the Newcastle-Ottawa Scale (NOS), which is designed for evaluating the quality of non-randomised studies, particularly cohort and case-control studies [28]. Longitudinal analyses from Randomised Controlled Trials (RCTs) were analysed as prospective cohort studies, and the NOS was applied to these studies accordingly to ensure consistent quality assessment across study designs. In the NOS, a ‘star system’ [28] has been developed in which a study is judged on the three main areas below:

1. Selection of participants (4 items): Evaluates how participants were chosen, whether inclusion/exclusion criteria were appropriate, and if groups were comparably selected.
2. Comparability of groups (1 item): Assesses whether the groups being compared are comparable on certain characteristics or controlling for confounding factors.
3. Outcome assessment (3 items): Looks at how outcomes were measured and whether the assessment methods were reliable and consistent.

The NOS for cohort studies was operationalised to fit our research question (see Appendix 4). Two review authors independently assessed the risk of bias in included studies (TS, HG).

Disagreements between the two assessors were resolved through discussion with a third review author (BLE). Certainty of evidence for each outcome was assessed using the Grading of Recommendations Assessment, Development and Evaluation (GRADE) system [29]. We used the GRADE approach adapted for narrative synthesis and further adapted to the purposes of our review [30]. NOS scores were interpreted to inform judgments about the overall risk of bias in the GRADE assessment.

### Data synthesis

A narrative synthesis was conducted informed by the Economic and Social Research Council guidance [31]. The characteristics of each included study were summarised, and findings were reported separately for each outcome: depression, suicidal ideation, anxiety and mixed outcomes. We classified studies by outcome rather than by population in order to reduce heterogeneity and facilitate a potential meta-analysis. However, this was deemed not feasible due to substantial variation in outcomes (e.g., included symptom severity, recurrence, service use), measured using different instruments and scales (e.g., Patient Health Questionnaire-9 (PHQ-9), the Center for Epidemiologic Studies Depression Scale (CES-D-10), the Composite International Diagnostic Interview (CIDI), electronic health record data) and statistical reporting. While most studies used adjusted models, the covariates included differed considerably, affecting the interpretation and pooling of effect sizes.

Based on study characteristics and following team discussions, we distinguished three settings:

A. General population samples (which may or may not include individuals receiving any form of care)
B. Populations recruited from mental health services (e.g., community mental health teams, but without a clearly defined specific intervention being delivered across the sample)
C. Populations receiving a specific clinical treatment (e.g., RCTs or other intervention studies).

We examined whether the association between loneliness and CMD outcomes varied within (and between) these settings.

### Ethics and data availability

This is a systematic review of previously reported studies in the public domain and requires no ethical approval or additional consent from participants.

## Results

### Study Selection

The bibliographic database search yielded 9086 citations: 7667 from the original search and 1419 from the updated search. Another 31 records were identified through Google Scholar and citation tracking. After removing duplicates, 7110 titles and abstracts were screened, and 623 full-text articles were assessed for eligibility. Seventeen studies met the inclusion criteria [32–48]. The study selection process, including reasons for exclusion, is detailed in the PRISMA flow diagram [25] (Fig. 1).

**Figure 1.**
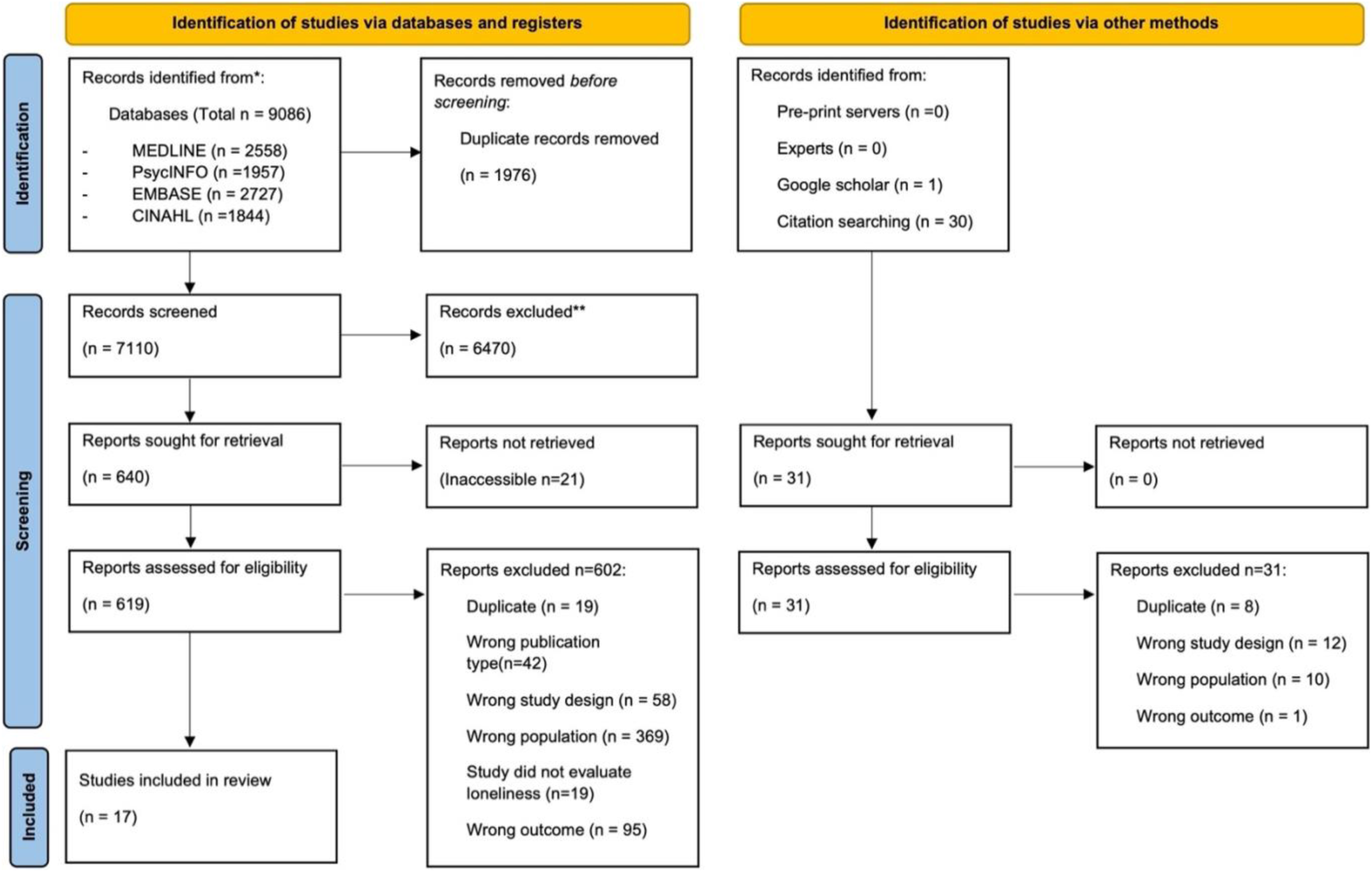
PRISMA flow diagram

Six additional studies were initially considered for inclusion as they examined associations between loneliness and CMD outcomes [49–54]. However, during the data extraction phase, it became evident that these associations were cross-sectional, not longitudinal, and thus did not meet the inclusion criteria. A summary of these ‘near misses’ is provided in Appendix 6.

### Study characteristics

The characteristics of the 17 included studies are presented in Table 1. All included studies were peer-reviewed journal articles published between 2010 and 2024. Sixteen studies were published in English, and one study (Ortiz et al. [40]) was published in Spanish. The studies were conducted in the Netherlands (n=8), the United States of America (USA) (n=3), Canada (n=1), Australia (n=1), Spain (n=1), Brazil (n=1), Mexico (n=1) and the United Kingdom (UK) (n=1). All included studies were longitudinal in design: fifteen were observational and two were intervention trials.

**Table 1.**
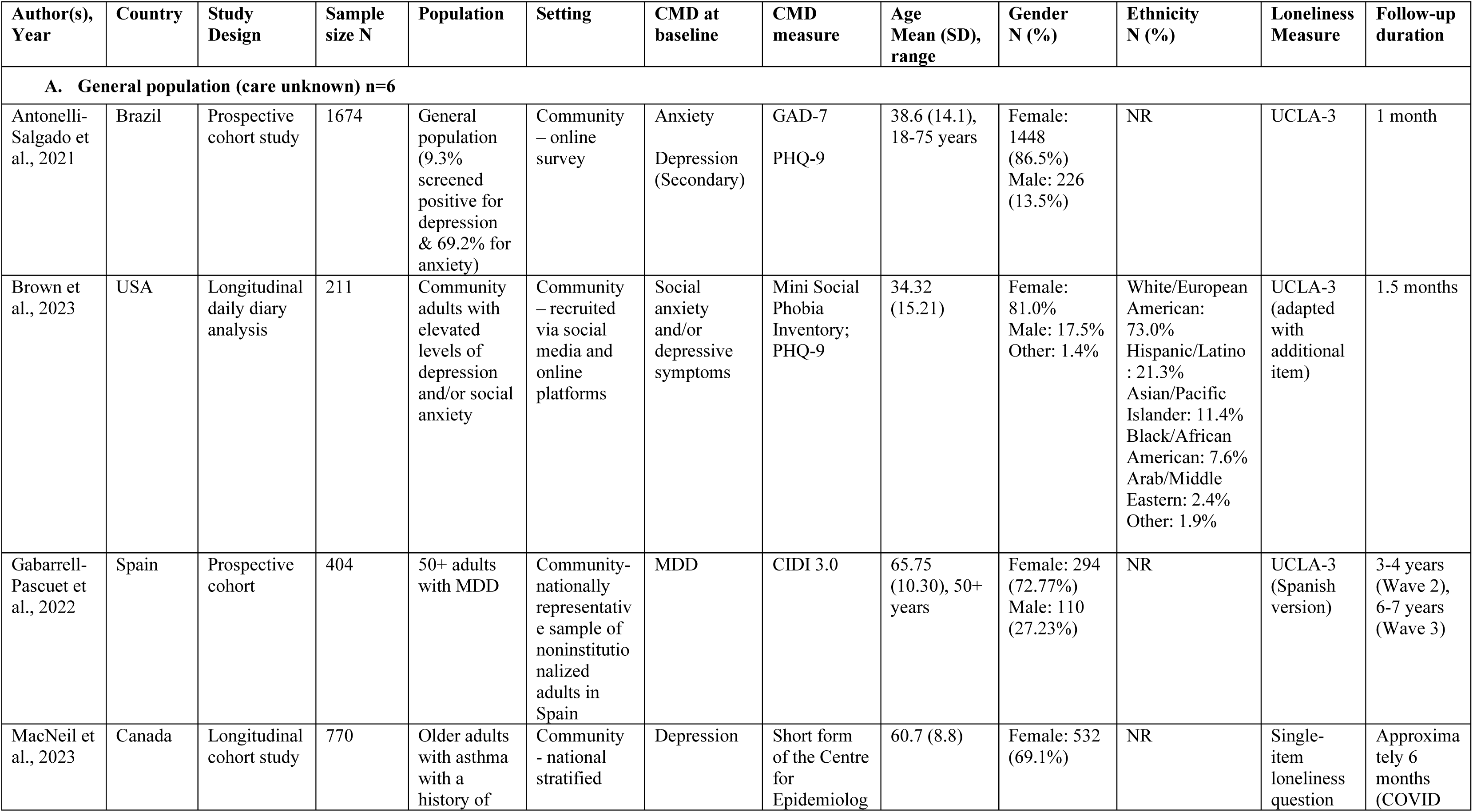
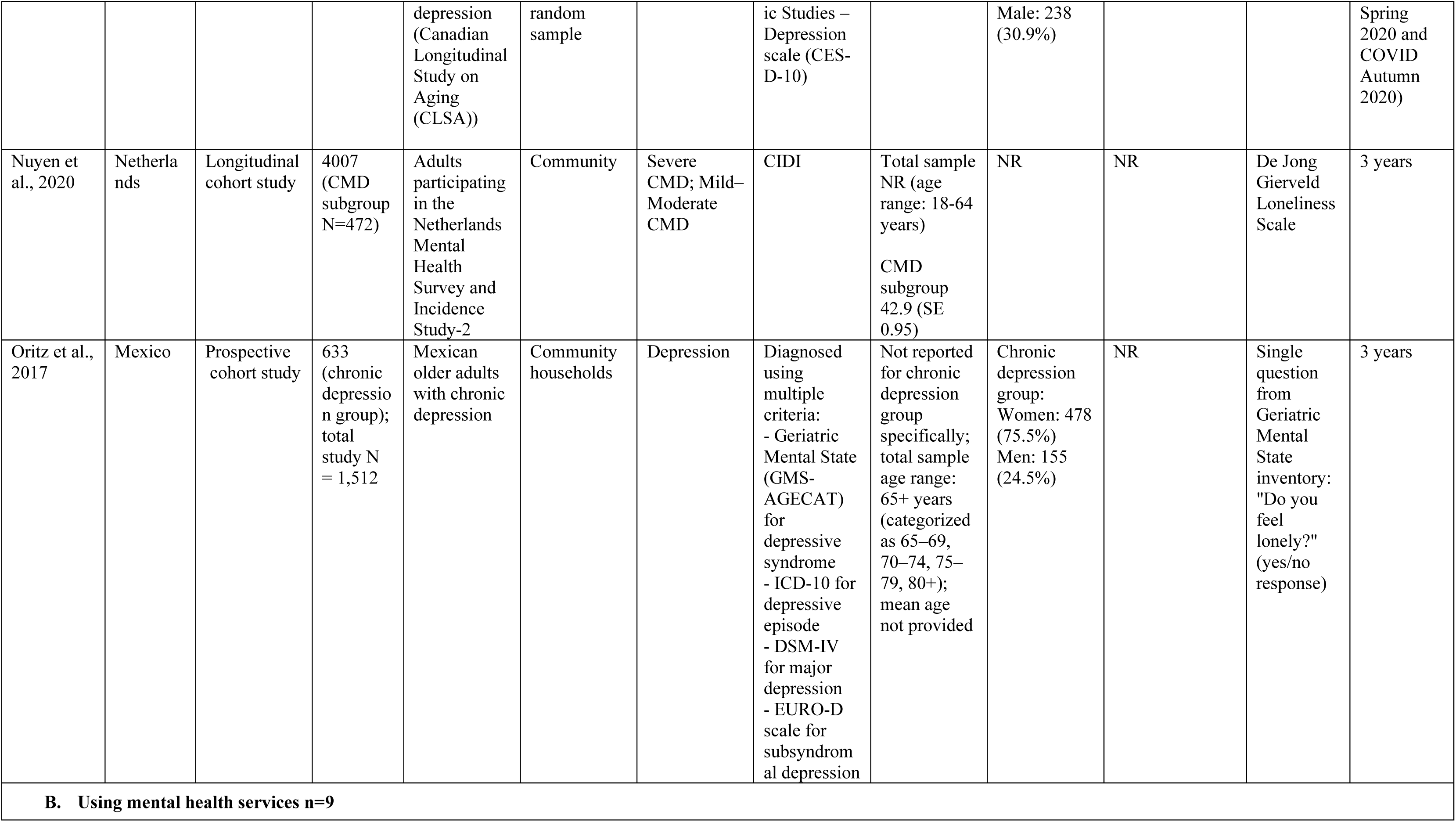
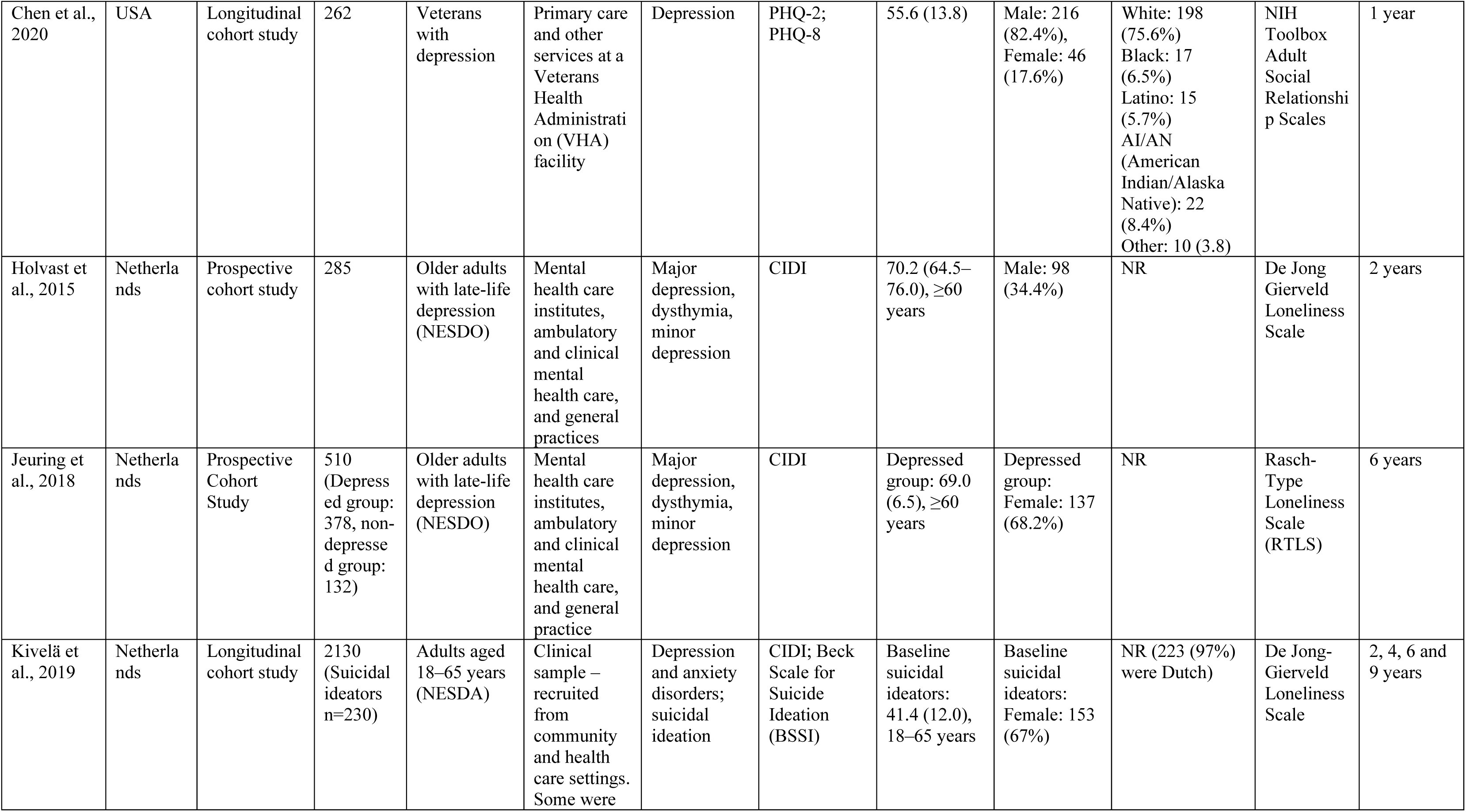
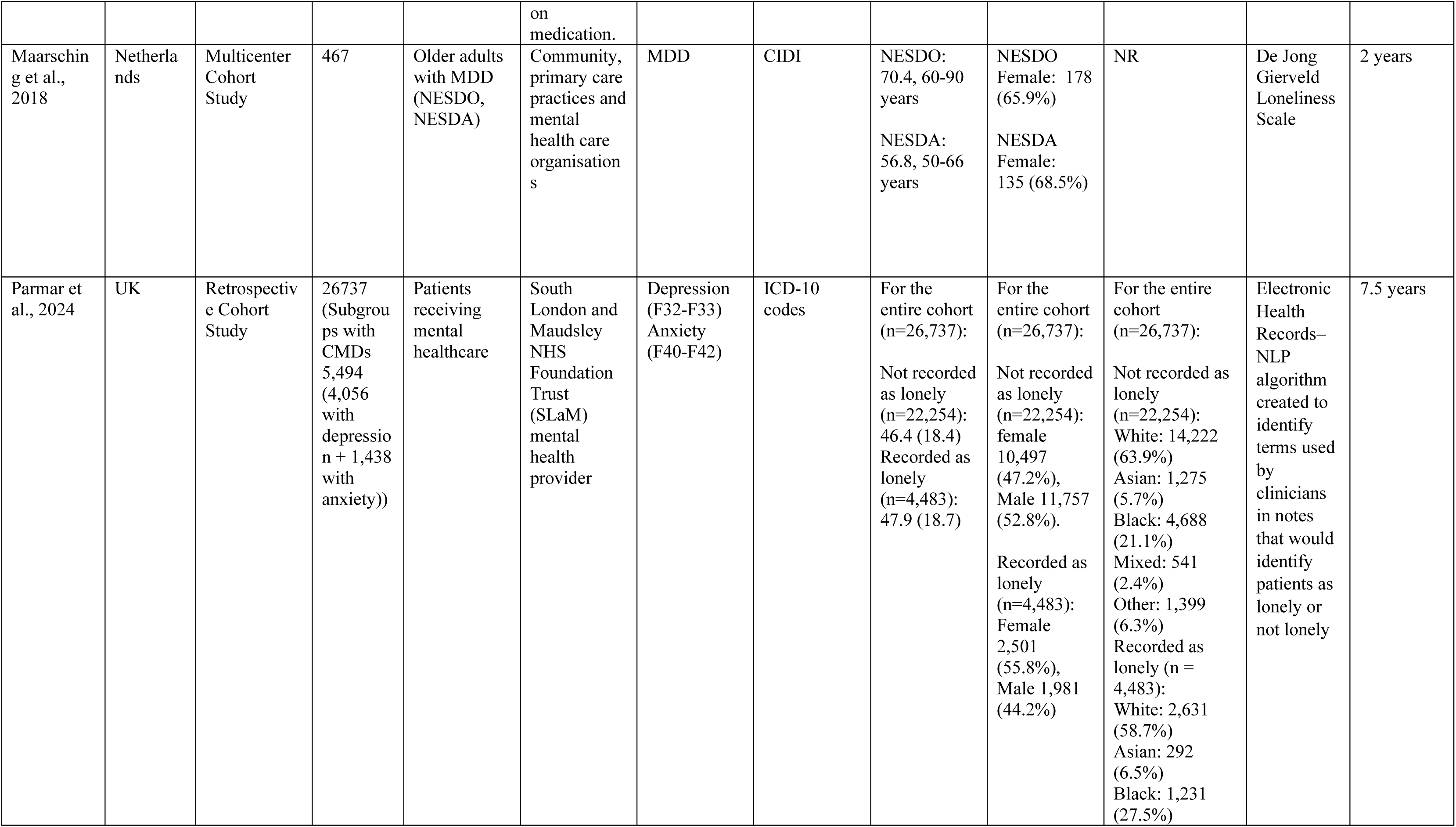
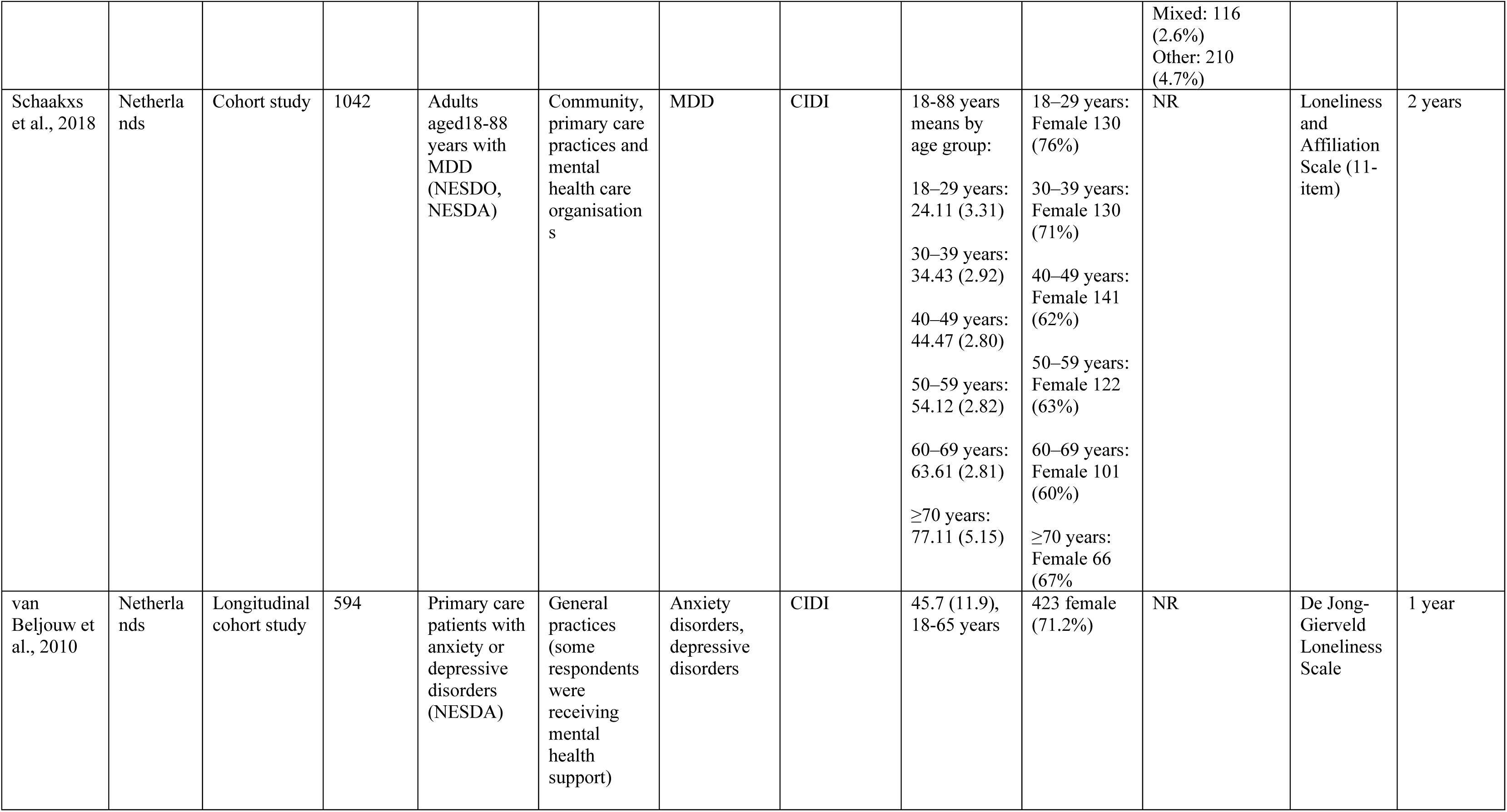
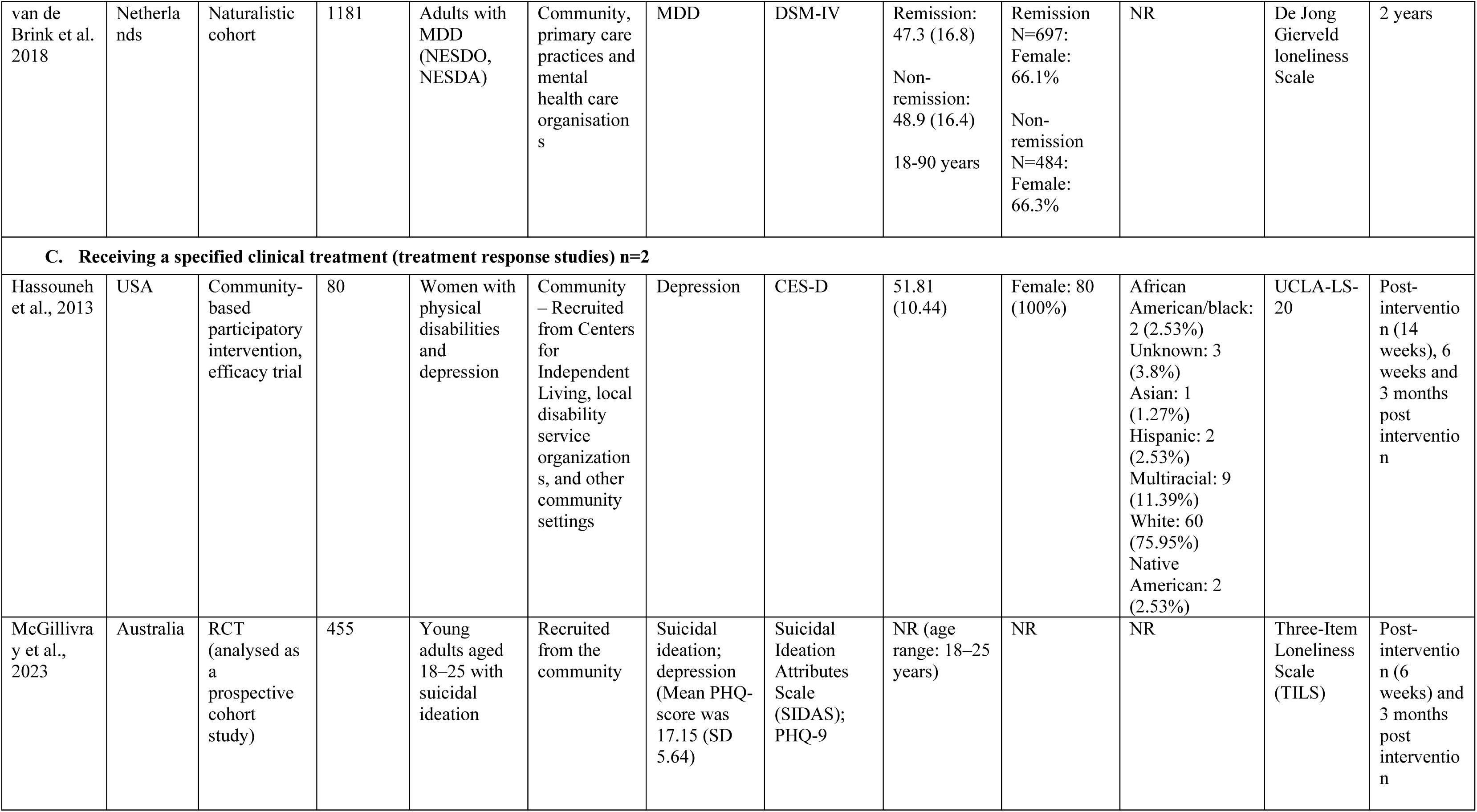
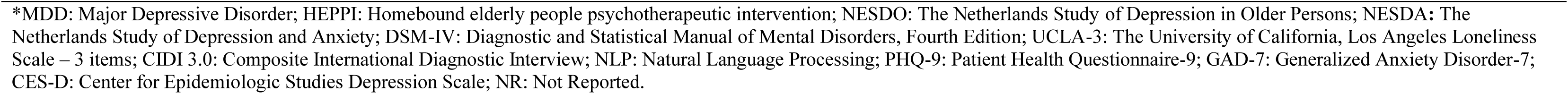
Characteristics of included studies N=17.

Populations by specific diagnosis varied, with eleven studies focusing on individuals with depression [32–42], four on mixed depression and anxiety [43–46], one on anxiety [47] and one on CMDs more broadly [48]. Many focused on older adults, particularly those analysing data from the Netherlands-based NESDA and NESDO cohorts [35–38, 40]. Sample sizes ranged from 80 to 26,737 participants (CMD subgroups: 80 to 5494).

Most studies assessed loneliness using validated scales, including the UCLA Loneliness scale and the De Jong Gierveld Loneliness Scale. Two studies used a single-item question [37, 40] and another used Natural Language Processing (NLP) algorithms to capture language in health records indicative of loneliness and distinguish lonely and not lonely patients [45]. Follow-up durations varied: five studies had follow-ups of less than one year, six ranged from one to two years, and six followed participants for over two years. Of the 17 included studies, two examined loneliness as a mediator of CMD outcomes [33, 43].

Findings are presented by outcome: depression, suicidal ideation, anxiety, and mixed CMD outcomes. Four studies contributed results across multiple outcome categories and are referenced in more than one section [32, 39, 45, 46]. A summary of findings by outcome and the direction of associations is presented in Fig. 2. Three treatment settings were distinguished: A. General population (care status unknown) (n= 6); B. Using mental health services (n=9); C. Receiving a specific clinical treatment (n=2).

**Figure 2.**
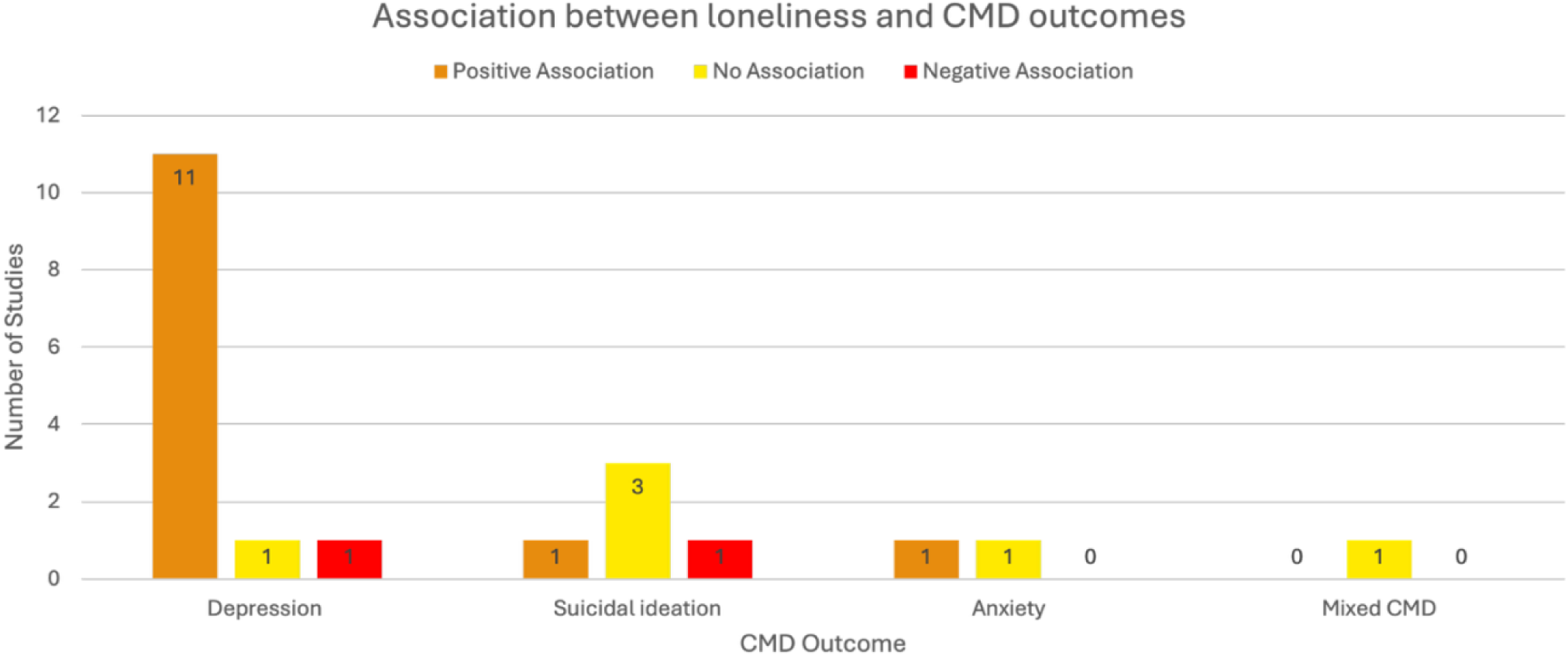
Summary of findings by outcome

#### Methodological quality

Regarding quality assessment, 13 studies (76%) were rated as high quality, and 4 studies (24%) were rated as moderate quality according to the NOS criteria. Common limitations included high attrition rates, lack of detail on follow-up procedures, limited adjustment for confounders and inadequate description of comparison groups. GRADE certainty of evidence ratings were high for depression outcomes, low for suicidal ideation, and very low for anxiety and mixed CMD outcomes. Further details of the quality assessments and GRADE ratings are provided in Appendix 4 and Appendix 5.

### Loneliness and Depression Outcomes

Thirteen studies examined the association between baseline loneliness and depression outcomes at follow-up (see Table 2) [32–42, 45–46]. Of these, eleven studies included samples of individuals with depression only [32–42], while two studies involved mixed samples of participants with depression and anxiety [45–46]. Studies were categorised by setting: general population (care status unknown, n=3), mental health services (n=8), and specific clinical treatment (n=2). There was no observed relationship between setting and statistical significance of findings.

**Table 2.**
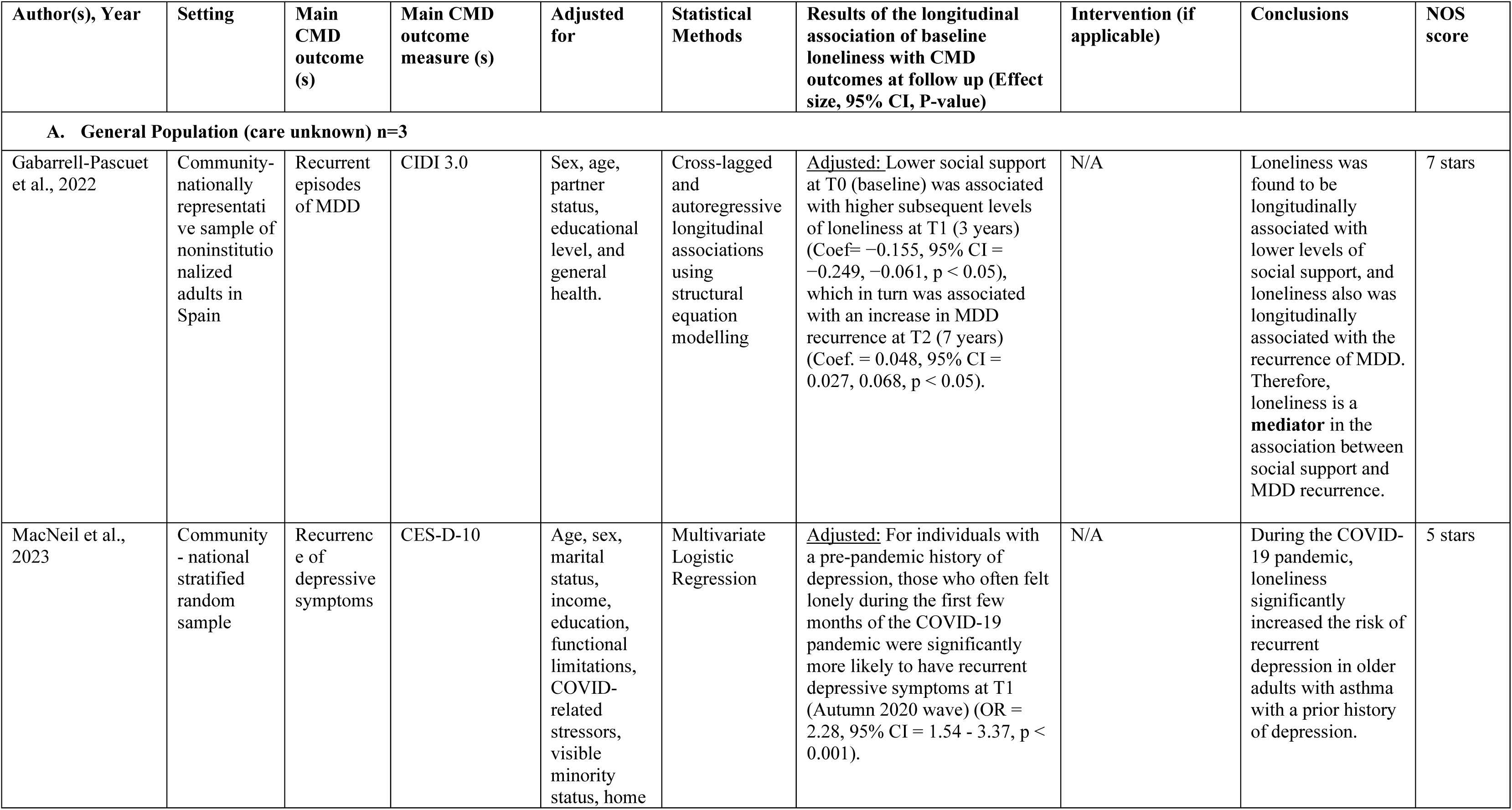
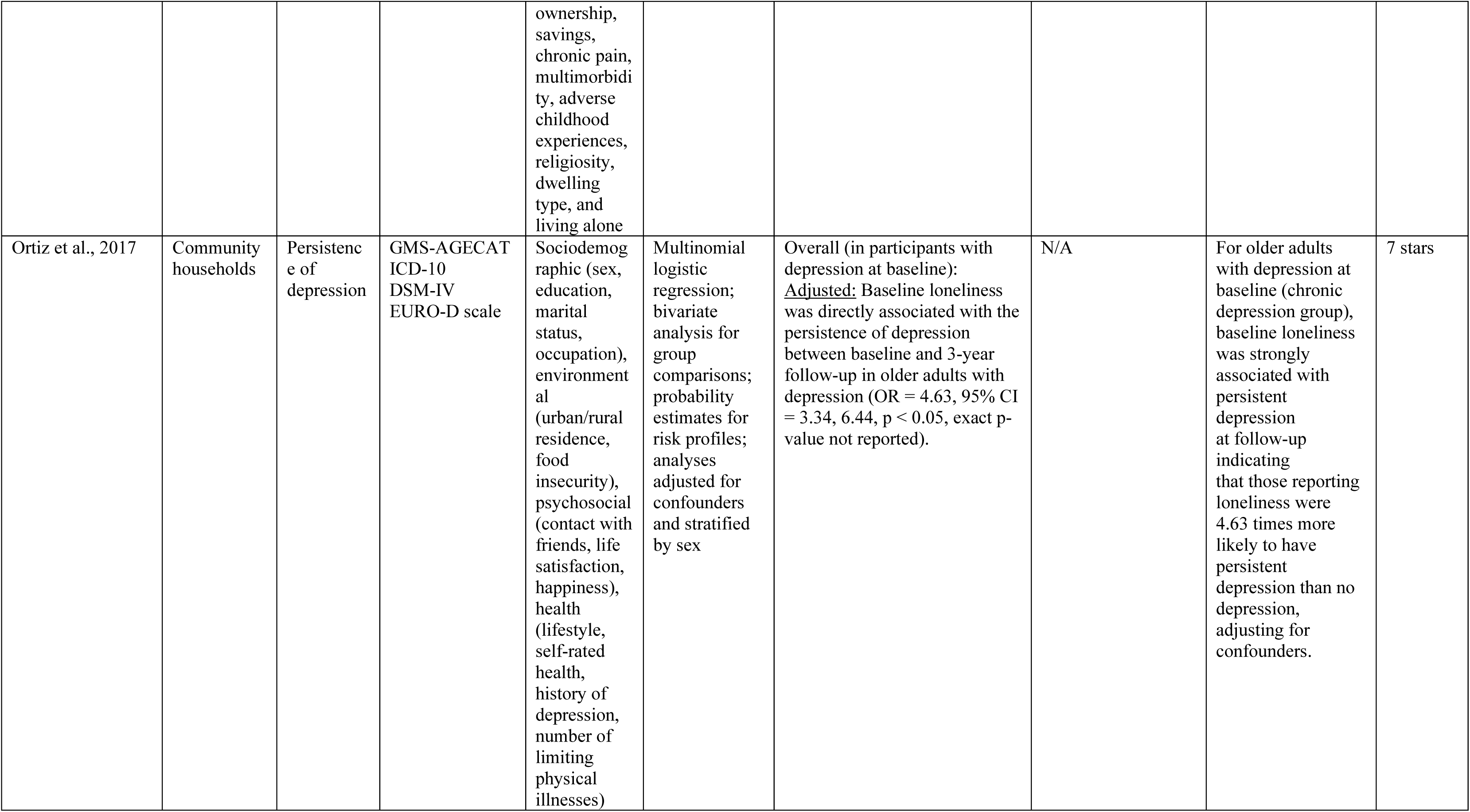
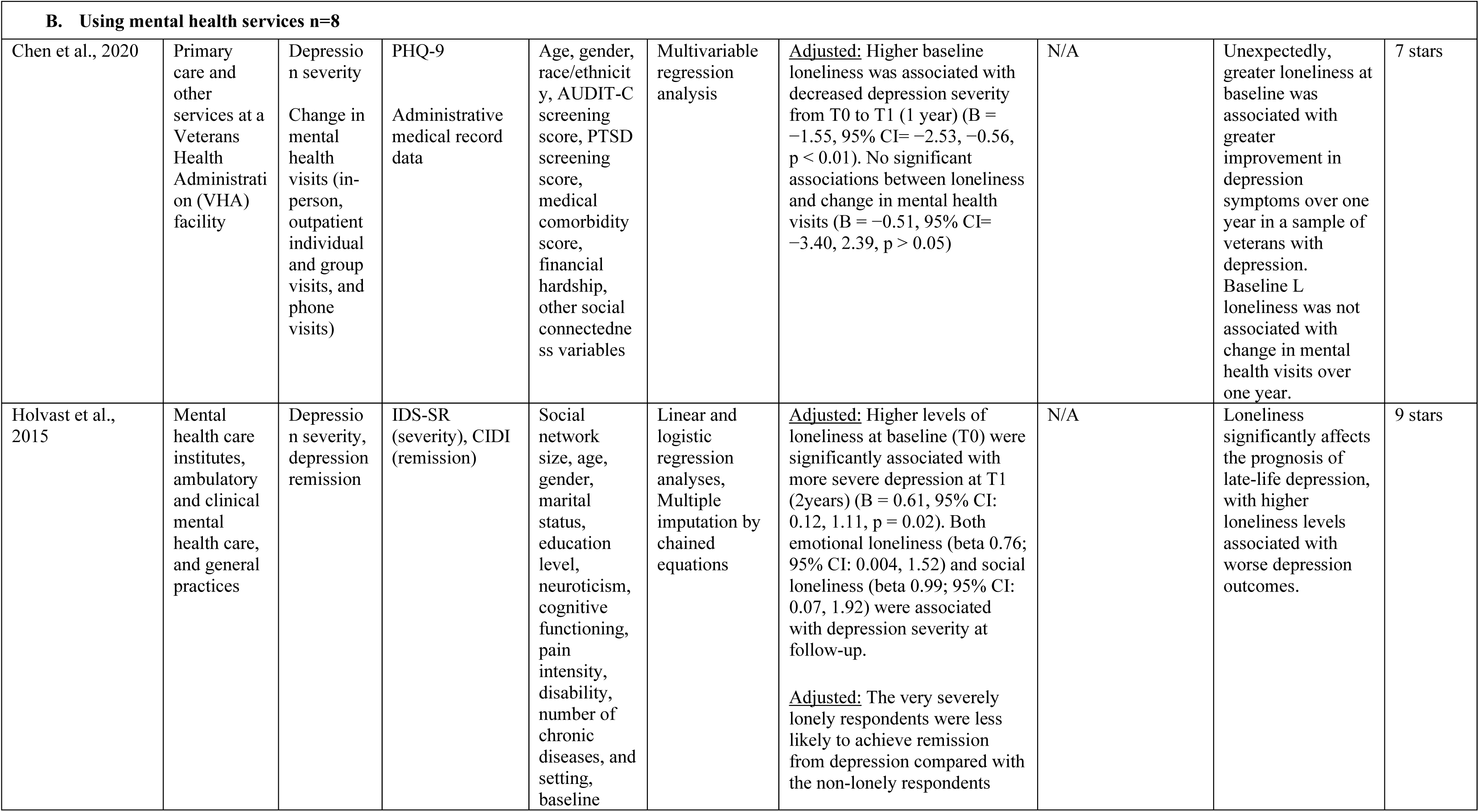
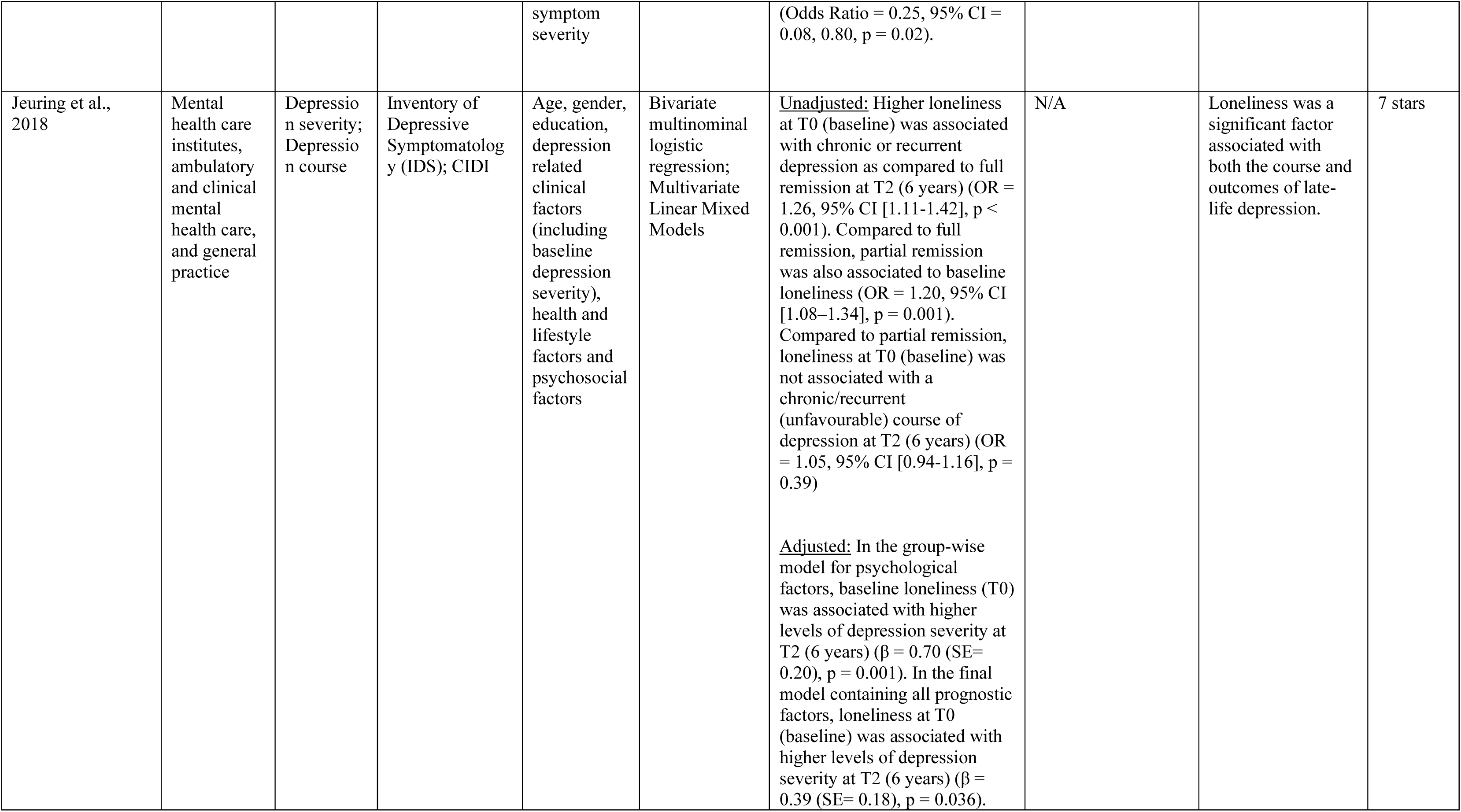
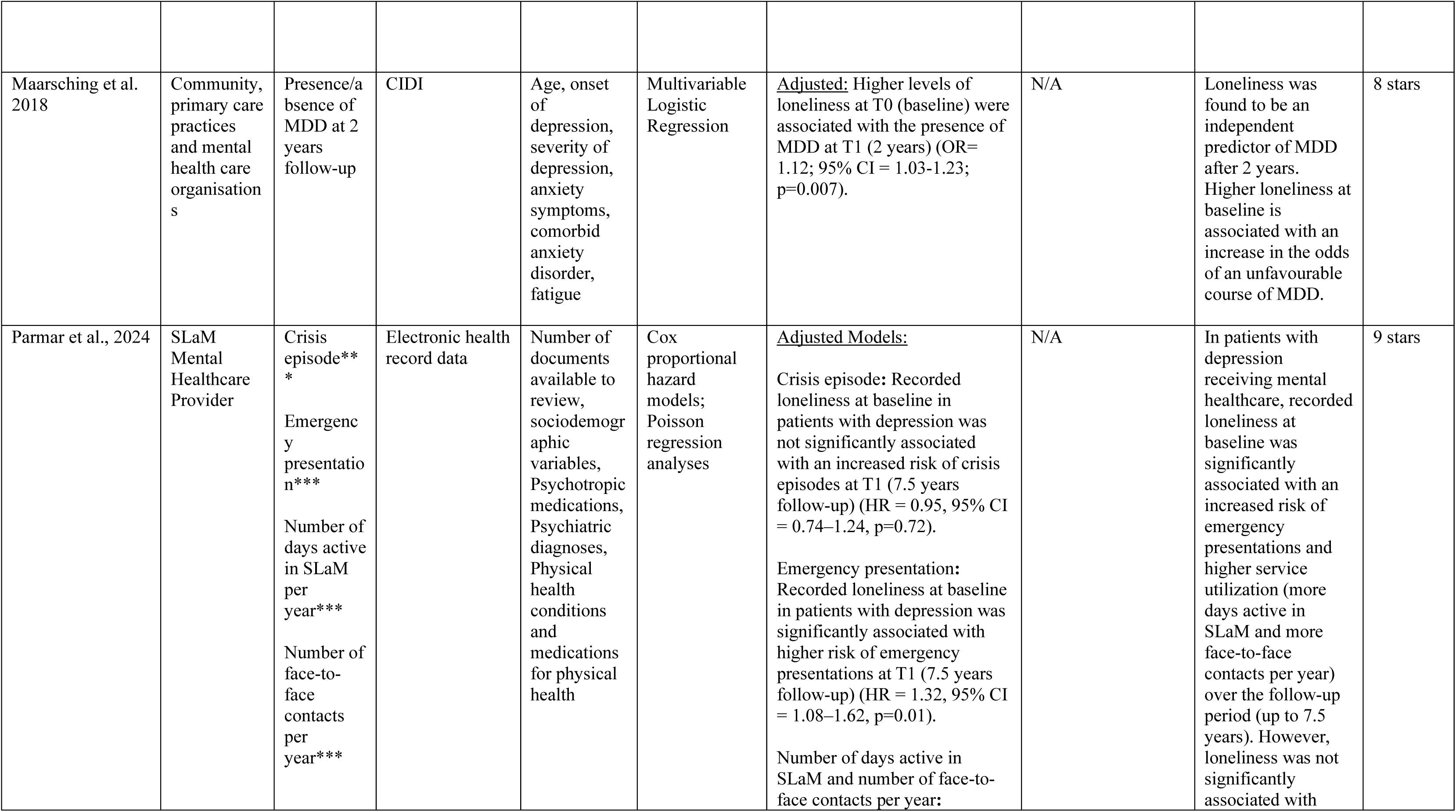
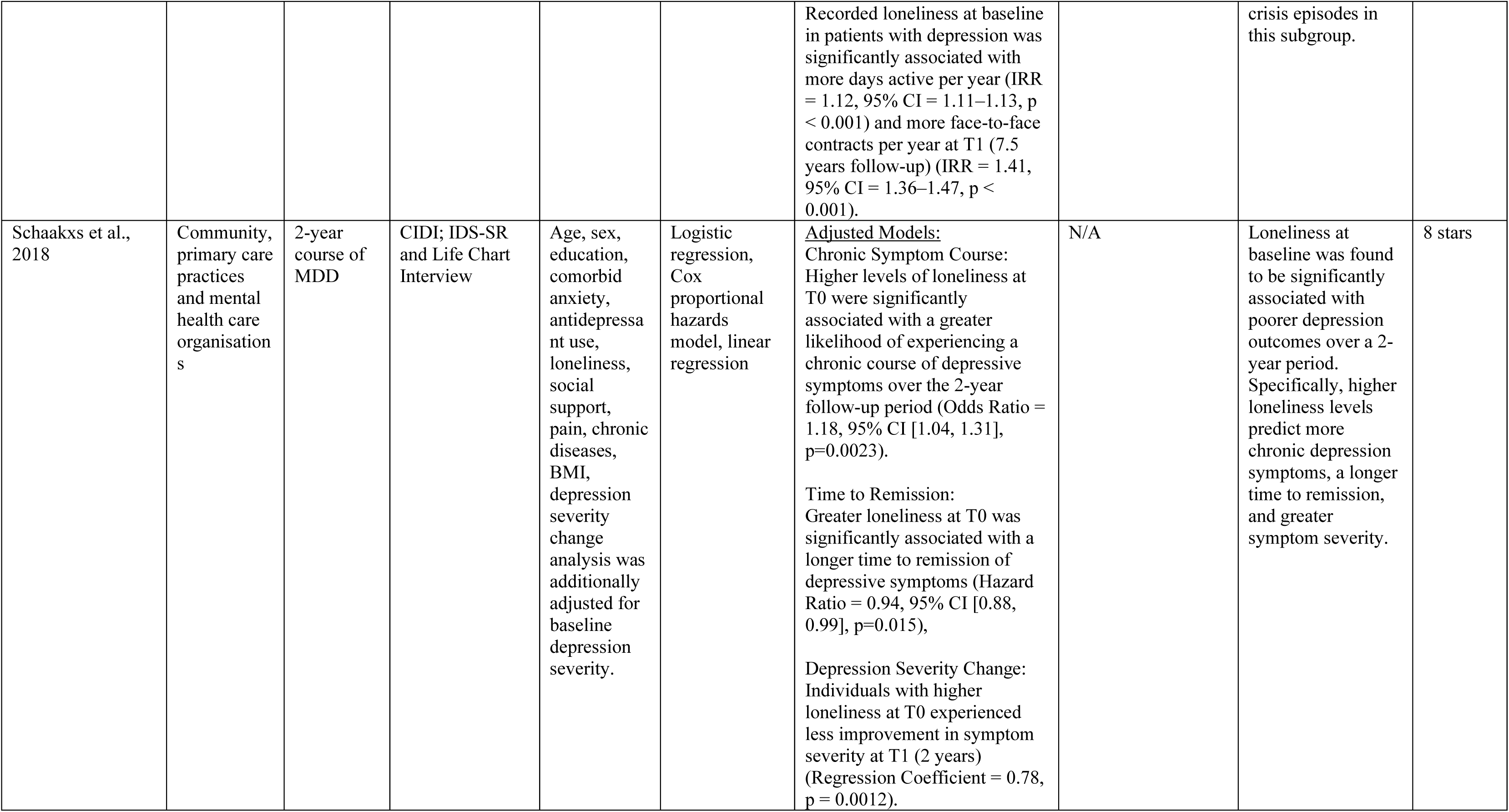
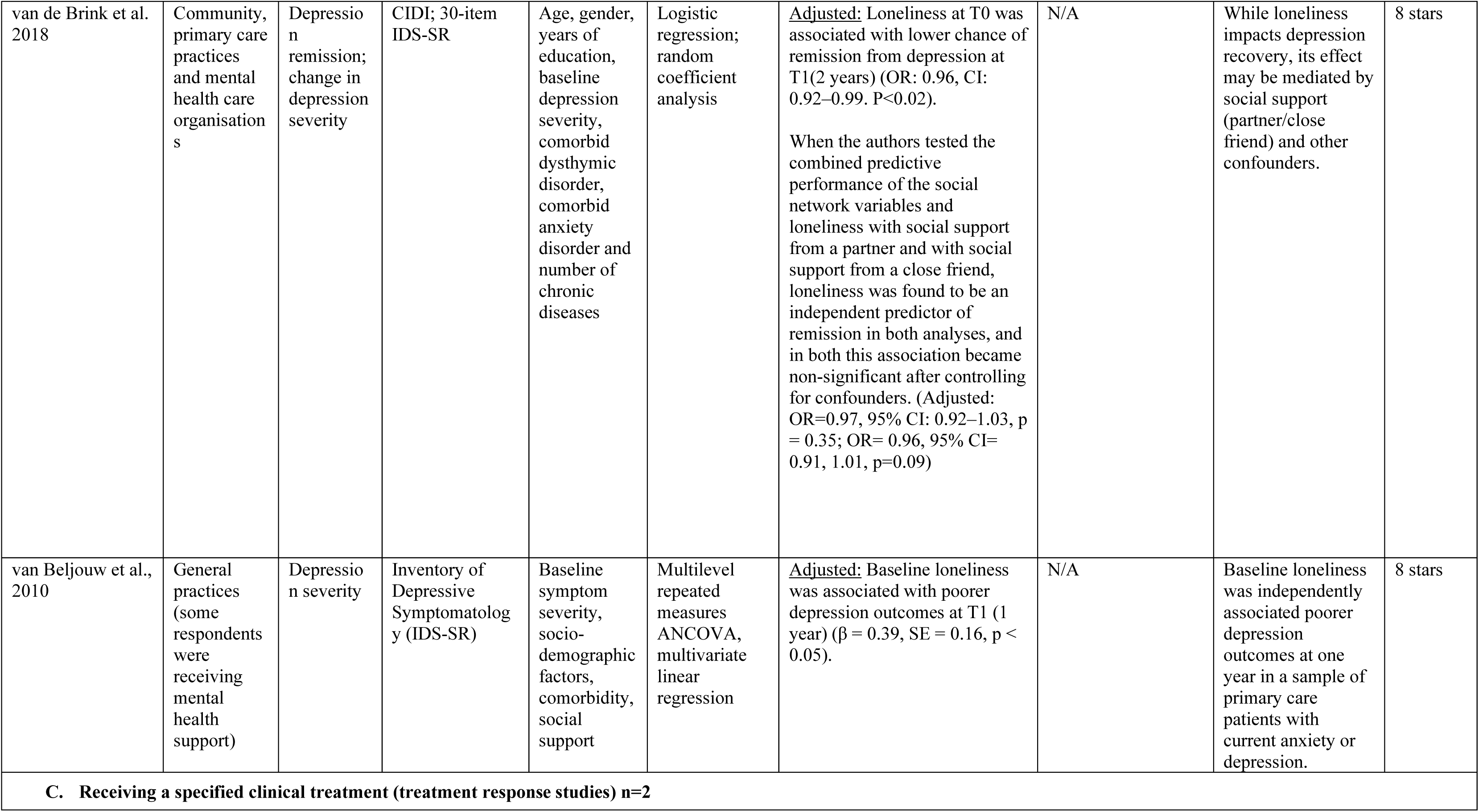
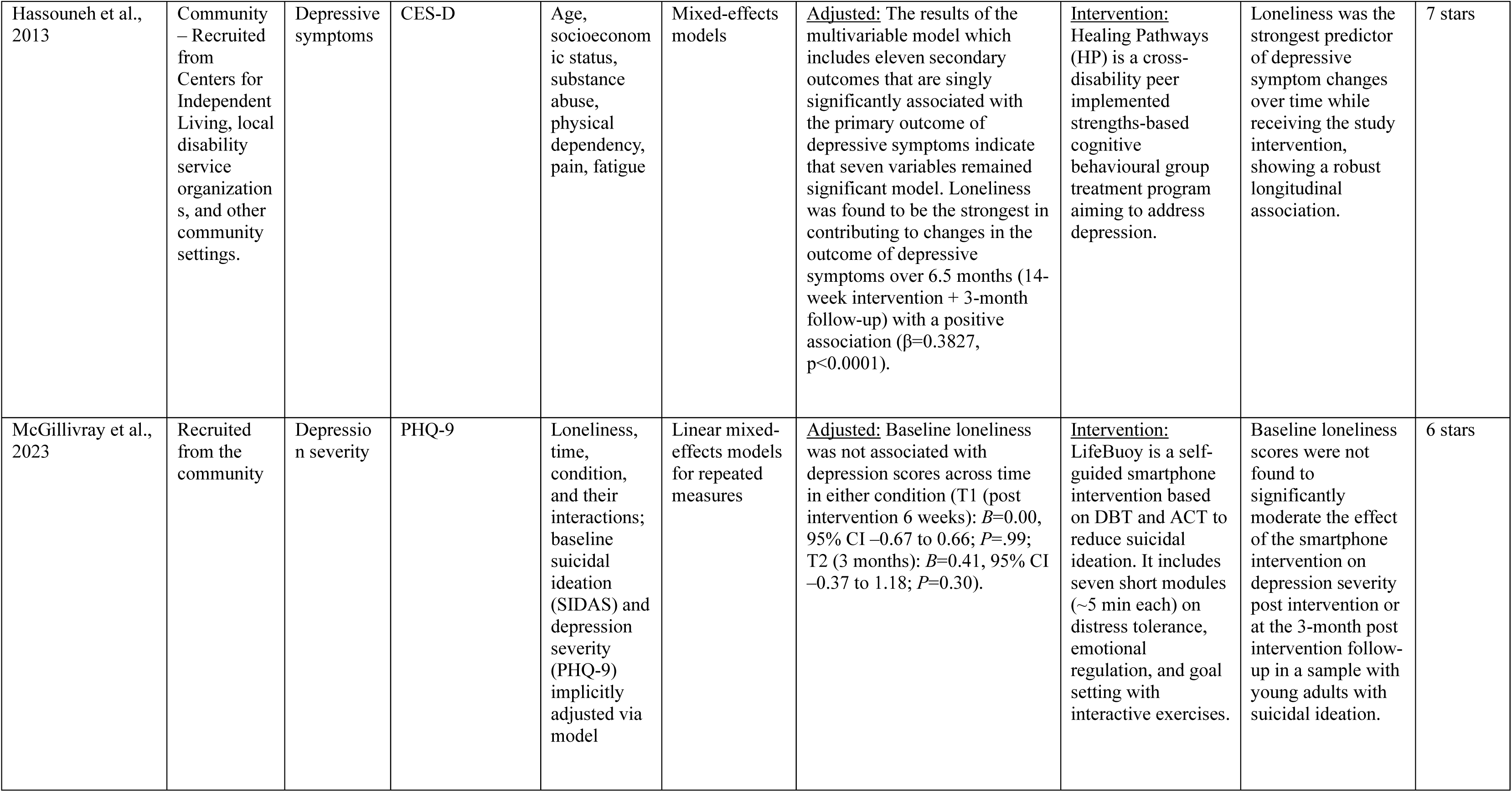
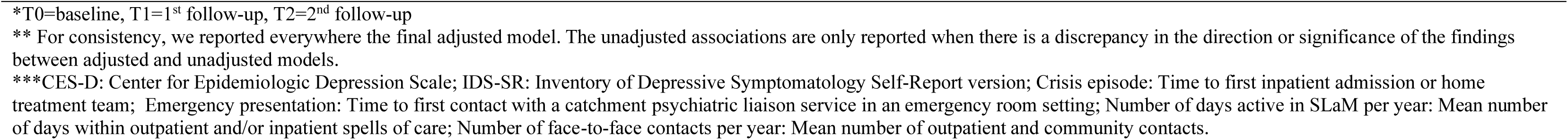
Study findings for depression outcomes N=13.

Depression-related outcomes included depression severity, course/remission, recurrence and service utilisation. Symptom-based outcomes were assessed using validated measures such as PHQ-9 [55], CES-D-10 [56], and CIDI [57]. Service utilisation outcomes were assessed using electronic health records or administrative data.

#### Positive associations

Eleven of the 13 studies (85%) found that greater loneliness at baseline was associated with worse depression outcomes at follow-up [33–38, 40–42,45–46]. Of these, nine studies reported worse clinical depression outcomes, including greater symptom severity, reduced remission, chronicity, or persistence of depression at follow-up. These findings were from both observational and intervention studies. Effect sizes ranged from weak to strong (ORs = 1.12–4.63; β = 0.38–0.78), with four studies [35, 37, 40, 41] reporting moderate to strong effects. One study [45] reported increased service utilisation, including emergency presentations (HR = 1.32, 95% CI [1.08–1.62], p = 0.01), days active in care per year (IRR = 1.12, 95% CI [1.11–1.13], p < 0.001), and face-to-face contacts per year (IRR = 1.41, 95% CI [1.36–1.47], p < 0.001), but no statistically significant association with crisis episodes (HR = 0.95, 95% CI [0.74–1.24], p = 0.72)

Only one study investigated mediators in this outcome category and identified loneliness as a mediator in the pathway between low social support and Major Depressive Disorder (MDD) recurrence, using a longitudinal structural equation model [33]. This finding further supports loneliness as a risk factor for poorer depression outcomes, even when not examined as a direct predictor. All studies employed adjusted models, indicating robust associations independent of confounding.

#### Negative or null associations

Two of the thirteen studies (15%) found a negative or null association between loneliness and depression outcomes. Chen et al. [32] reported that higher baseline loneliness was associated with greater improvement in depressive symptoms over one year in a sample of veterans (B = −1.55, 95% CI [−2.53, −0.56], p < 0.01). The authors suggested this unexpected finding may reflect population-specific effects or regression to the mean. The study also found no association between loneliness and mental health visits at follow up (B = −0.51, 95% CI [ −3.40, 2.39], p > 0.05). McGillivray et al. [39], an RCT of a digital self-guided Dialectical Behaviour Therapy (DBT)/ Acceptance and Commitment Therapy (ACT) intervention for young adults with suicidal ideation, found no significant association between baseline loneliness and depression symptom outcomes either at post-treatment (B = 0.00, 95% CI [–0.67, 0.66], p = 0.99) or at 3-month follow-up (B = 0.41, 95% CI [–0.37, 1.18], p = 0.30). Loneliness did not moderate depression trajectories across conditions.

#### Intervention studies

Among the two intervention studies, findings were mixed. Hassouneh et al. [34], a strengths-based, peer-delivered Cognitive Behavioural Therapy (CBT) program for women with physical disabilities, found that baseline loneliness was the strongest predictor of change in depressive symptoms over time. In contrast, McGillivray et al. [39], as described above, found no relationship between loneliness and depression outcomes or intervention response.

Taken together, these findings suggest that the association between loneliness and subsequent depression was consistent across populations and study designs. The GRADE certainty of evidence for the thirteen studies reporting on the associations between loneliness and depression outcomes was rated as high. According to NOS ratings, eleven studies (85%) were of high methodological quality (7–9 stars), and 2 studies (15%) were of moderate quality (5-6 stars).

### Loneliness and Suicidal Ideation

Five studies examined the relationship between baseline loneliness and suicidal ideation at follow-up (see Table 3) [32, 39, 43, 44, 47]. In all studies, suicidal ideation was measured using validated measures (e.g., Beck Depression Inventory (BDI-II) suicide item, Beck Scale for suicide ideation) except for one study where a single self-report question was used [47]. Two studies were conducted in general population samples, two involved individuals using mental health services, and one examined a sample receiving a specific clinical treatment.

**Table 3.** Study findings for suicidal ideation outcomes N=5.

| Author(s), Year | Setting | Main CMD outcome (s) | Main CMD outcome measure (s) | Adjusted for | Statistical Methods | Results of the longitudinal association of baseline loneliness with CMD outcomes at follow up (Effect size, 95% CI, P-value) | Intervention (if applicable) | Conclusions | NOS score |
| --- | --- | --- | --- | --- | --- | --- | --- | --- | --- |
| **A. General Population (care unknown) n=2** |  |  |  |  |  |  |  |  |  |
| Antonelli-Salgado 2021 | Community – online survey | Suicidal ideation | Self-report to a single question | Age, gender, sexual orientation, marital status, income, geographical area, education, profession, unemployment, financial crisis during the pandemic, quality of family relationships, quality of friend relationships, religion, meditation, sleep quality, physical activity, childhood trauma, previous suicide attempt, family history of suicide, GAD-7, PHQ-9, benzodiazepines use and AUDIT-C | Multinomial logistic regression analysis | <u>Adjusted:</u> Baseline loneliness was directly associated with the incidence of suicidal ideation between T0 and T1 (1 month) (OR= 2.12, 95% CI = 1.06, 4.24, p=0.033). | N/A | Loneliness was consistently associated with the incidence of suicidal ideation in a one-month follow-up during the initial stage of the COVID-19 pandemic in a general population sample with anxiety symptoms. | 6 stars |
| Brown et al., 2023 | Community – recruited via social media and online platforms | Suicidal ideation | BDI-II Suicide Item | Baseline suicidal ideation and social anxiety | Mediation Analysis with bootstrapping | <u>Adjusted:</u> Loneliness at T1 (14-day daily diary period) did not predict suicidal ideation at T2 (1.5 month) when controlling for baseline social anxiety and baseline suicidal ideation ( $\beta = 0.08, p = 0.25$ ). Loneliness did not mediate the social anxiety–SI relationship (indirect effect: $\beta = 0.03$ , Bootstrap CIs $[-0.02, 0.08]$ ). | N/A | Loneliness did not significantly mediate the relationship between social anxiety and suicidal ideation at 1.5-month follow-up in a sample of community adults with elevated depression and/or social anxiety. | 5 stars |
| **B. Using mental health services n=2** |  |  |  |  |  |  |  |  |  |
| Chen et al., 2020 | Primary care and other services at a Veterans Health Administration (VHA) facility | Change in Suicidal ideation | PHQ-9 (item 9) | Age, gender, race/ethnicity, AUDIT-C screening score, PTSD screening score, medical comorbidity score, financial hardship, other social connectedness variables | Multivariable regression analysis | <u>Adjusted:</u> Higher baseline loneliness was associated with decreased suicidal ideation from T0 to T1 (1 year) ($B = -0.21$, 95% CI [not reported], $p < 0.05$). | N/A | Unexpectedly, higher levels of baseline loneliness were associated with greater improvement in suicidal ideation over one year in a sample of veterans with depression. The authors suggest that these results contrast with previous studies and may be due to population-specific characteristics and regression to the mean. | 7 stars |
| Kivelä et al., 2019 | Clinical sample – recruited from community and health care settings. Some were on medication. | Persistence of suicidal ideation | Beck Scale for Suicide Ideation (BSSI) | Sociodemographic (age, gender, marital status, education, employment), clinical factors (depression, anxiety, comorbidity, symptom severity, medication use, suicide attempt history), baseline predictors (insomnia symptoms, hopelessness, loneliness, BPD traits, childhood trauma, negative life events) | Linear mixed-effects growth models, logistic regression | <u>Adjusted:</u> Baseline loneliness was not significantly associated with persistent suicidal ideation over 9 years ($OR = 1.03$, 95% CI = $0.88-1.20$, $p > 0.05$). | N/A | Loneliness did not predict the course or persistence of suicidal ideation in a sample of individuals with suicidal ideation. | 7 stars |
| **C. Receiving a specified clinical treatment (treatment response studies) n=1** |  |  |  |  |  |  |  |  |  |
| McGillivray et al., 2023 | Recruited from the community | Suicidal ideation | Suicidal Ideation Attributes Scale (SIDAS) | Loneliness, time, condition, and their interactions; baseline suicidal ideation (SIDAS) and depression severity (PHQ-9) implicitly adjusted via model | Linear mixed-effects models for repeated measures | <u>Adjusted:</u> Baseline loneliness was not associated with suicidal ideation scores across time in either condition ($B=1.10$, 95% CI $-0.25$ to $2.46$; $P=0.11$ at T1 (post intervention 6 weeks); $B=0.43$, 95% CI $-1.25$ to $2.12$; $P=0.61$ at T2 (3 months post intervention follow-up)). | <u>Intervention:</u> LifeBuoy is a self-guided smartphone intervention based on DBT and ACT to reduce suicidal ideation. It includes seven short modules (~5 min each) on distress tolerance, emotional regulation, and goal setting with interactive exercises. | Baseline loneliness scores were not found to significantly moderate the effect of the smartphone intervention on suicidal ideation post intervention or at the 3-month post intervention follow-up in a sample with young adults with suicidal ideation. | 6 stars |

\*T0=baseline, T1=1<sup>st</sup> follow-up, T2=2<sup>nd</sup> follow-up
\*\* For consistency, we reported everywhere the final adjusted model. The unadjusted associations are only reported when there is a discrepancy in the direction or significance of the findings between adjusted and unadjusted models.

Only one study of the five found a positive association, indicating that greater loneliness at baseline was associated with higher incidence of suicidal ideation in a one-month follow-up during the COVID-19 pandemic in a general population sample with anxiety symptoms (OR = 2.12, 95% CI [1.06, 4.24], p = 0.033) [47].

In contrast, two studies reported no significant association between loneliness and subsequent suicidal ideation: one in a clinical sample of adults with depression and anxiety [44], and another in young adults with suicidal ideation [39] (p > 0.05). The latter evaluated a digital self-guided intervention for young adults with suicidal ideation and found no significant effect of baseline loneliness on suicidal ideation outcomes, either at post-treatment (B = 1.10, 95% CI [–0.25, 2.46], p = 0.11) or at three-month follow-up (B = 0.43, 95% CI [–1.25, 2.12], p = 0.61). These findings mirrored those reported in the previous section on depression outcomes, where loneliness was also not found to significantly predict depression severity in this trial.

Chen et al. [32] reported an inverse relationship: higher loneliness levels at baseline were associated with greater improvement in suicidal ideation over one year in a sample of veterans with depression (B = –0.21, p < 0.05). The authors suggested that this unexpected result may reflect regression to the mean or population-specific factors.

In the sole study in this category investigating mediators, loneliness did not significantly mediate the relationship between social anxiety and suicidal ideation at 1.5-month follow-up in a sample of community adults with elevated depression and/or social anxiety [43].

Taken together, the current evidence does not suggest a consistent association between loneliness and subsequent suicidal ideation among people with CMDs. The GRADE certainty of evidence was low, reflecting serious concerns about inconsistency in findings and imprecision due to small sample sizes. Three studies were of moderate methodological quality (5-6 stars) and two were of high quality (7 stars).

### Loneliness and Anxiety or Mixed CMD outcomes

Three studies examined the relationship between baseline loneliness and anxiety or mixed CMD outcomes at follow-up (see Table 4) [45, 46, 48]. Two studies focused specifically on anxiety [45, 46], while one investigated the severity of mixed CMD outcomes [48]. Outcomes included anxiety severity, service utilisation and CMD severity.

**Table 4.** Study findings on anxiety and mixed CMD outcomes N=3.

| Author(s), Year | Setting | Main CMD outcome (s) | Main CMD outcome measure (s) | Adjusted for | Statistical Methods | Results of the longitudinal association of baseline loneliness with CMD outcomes at follow up (Effect size, 95% CI, P-value) | Intervention (if applicable) | Conclusions | NOS score |
| --- | --- | --- | --- | --- | --- | --- | --- | --- | --- |
| **A. General Population (Care unknown) – mixed outcomes n=1** |  |  |  |  |  |  |  |  |  |
| Nuyen et al., 2020 | Community | Severity of CMD (mild/moderate vs severe) | CIDI | Gender, age, education, living situation, job status, and household income situation, negative life events, chronic physical disorders, BMI, perceived social support | Multivariate multinomial logistic regression | In the subgroup with CMD at baseline:<br><u>Adjusted:</u> For respondents with CMD at baseline, loneliness at T0 was not associated with persistent mild–moderate CMD at T1 (3 years) (Relative Risk Ratio [RRR] = 1.15, 95% CI [0.54, 2.47]).<br><u>Unadjusted:</u> For respondents with CMD at baseline, loneliness at T0 significantly predicted persistent severe CMD at follow-up (Relative Risk Ratio [RRR] = 2.48, 95% CI: 1.21–5.11, $p < 0.05$). | N/A | There was no significant relationship between loneliness and persistent mild–moderate CMD at follow-up across all models.<br>Loneliness predicted persistent severe CMD in unadjusted models, but social support attenuated this effect in adjusted models. | 8 stars |
|  |  |  |  |  |  | <u>Adjusted:</u> The association became non-significant after adjusting for perceived social support (Adjusted RRR = 1.80, 95% CI: 0.83-3.91). |  |  |  |
| **B. Using mental health services – anxiety n=2** |  |  |  |  |  |  |  |  |  |
| van Beljouw et al., 2010 | General practices (some respondents were receiving mental health support) | Anxiety symptom severity | Beck Anxiety Inventory (BAI) | Baseline symptom severity, socio-demographic factors, comorbidity, social support | Multilevel repeated measures ANCOVA, multivariate linear regression | <u>Unadjusted:</u> Baseline loneliness was significantly associated with poor anxiety outcomes at T1 (1 year) ($\beta = 0.40$, SE = 0.12, $p < 0.05$).<br><u>Adjusted:</u> Baseline loneliness was not significantly associated with poor anxiety outcomes at T1 (1 year) after adjustments ($\beta = -0.02$, SE = 0.10). |  | After adjustment, baseline loneliness was not associated with anxiety outcomes at 1 year follow-up. | 8 stars |
| Parmar et al., 2024 | SLaM Mental Healthcare Provider | Crisis episode***<br>Emergency presentation ***<br>Number of days active in SLaM per year*** | Electronic health record data | Number of documents available to review, sociodemographic variables, Psychotropic medications, Psychiatric diagnoses, Physical health conditions and medications for physical health | Cox proportional hazard models; Poisson regression analyses | <u>Adjusted Models:</u><br>Crisis episode: Recorded loneliness at baseline in patients with anxiety was not significantly associated with an increased risk of crisis episodes at T1 (7.5 years follow-up) (HR |  | Baseline loneliness in patients with anxiety was significantly associated only with increased face-to-face contacts per year over 7.5 years. No significant associations were found with crisis episodes, emergency presentations, or days | 9 stars |
| | | Number of face-to-face contacts per year*** | | | | = 1.11, 95% CI = 0.65–1.90, p = 0.70).<br>Emergency presentation:<br>Recorded loneliness at baseline in patients with anxiety was not significantly associated with higher risk of emergency presentations at T1 (7.5 years follow-up) (HR = 0.92, 95% CI = 0.62–1.37, p = 0.68).<br>Number of days active in SLaM per year:<br>Recorded loneliness at baseline in patients with anxiety was not significantly associated with the number of days active in SLaM per year at T1 (7.5 years follow-up) (IRR = 0.95, 95% CI = 0.88–1.02, p = 0.18).<br>Number of face-to-face contacts per year:<br>Recorded loneliness at baseline in patients with anxiety was significantly associated with an increased number of face-to-face contacts<br>per year at T1 (7.5<br>years follow-up) (IRR<br>= 1.24, 95% CI =<br>1.13–1.37, $p < 0.001$ ). | | active in SLaM. Loneliness may drive increased clinician engagement in anxiety patients, warranting further research into targeted interventions. | |

\*T0=baseline, T1=1<sup>st</sup> follow-up, T2=2<sup>nd</sup> follow-up
\*\* For consistency, we reported everywhere the final adjusted model. The unadjusted associations are only reported when there is a discrepancy in the direction or significance of the findings between adjusted and unadjusted models.
\*\*\* Crisis episode: Time to first inpatient admission or home treatment team; Emergency presentation: Time to first contact with a catchment psychiatric liaison service in an emergency room setting; Number of days active in SLAM per year: Mean number of days within outpatient and/or inpatient spells of care; Number of face-to-face contacts per year: Mean number of outpatient and community contacts.

Findings related to anxiety were limited and inconsistent. van Beljouw et al. [46] found no significant association between baseline loneliness and anxiety symptoms at one-year follow-up after adjustment (β = –0.02, SE = 0.10). Parmar et al. [45] reported mixed effects: loneliness was not associated with crisis episodes, emergency presentations, or days active in care, but was significantly associated with increased face-to-face contacts per year (IRR = 1.24, 95% CI [1.13–1.37], p < 0.001).

In the only study on mixed CMD outcomes, Nuyen et al. [48] examined mixed CMD severity in a general population sample of individuals with CMD at baseline. Loneliness at baseline did not significantly predict persistent mild-to-moderate CMD at three-year follow-up (Adjusted RRR = 1.15, 95% CI [0.54–2.47]), nor persistent severe CMD after controlling for perceived social support (Adjusted RRR = 1.80, 95% CI [0.83–3.91]). Although a significant unadjusted association was initially observed (RRR = 2.48, 95% CI [1.21–5.11]), this was attenuated in adjusted models.

All three studies were of high methodological quality (8-9 stars). However, the GRADE certainty of evidence for both anxiety and mixed CMD outcomes was rated as very low. This was due to serious concerns about inconsistency, imprecision (e.g., small number of studies and conflicting results), and potential publication bias.

## Discussion

### Summary of findings

This systematic review examined the longitudinal associations between baseline loneliness and follow-up mental health outcomes in individuals with CMDs. This review addressed an important gap in the literature regarding the prognostic role of loneliness in those already affected by CMDs, regardless of whether they receive treatment for their CMD. Seventeen studies were included, with the majority (77%) focusing on depression-related outcomes. Evidence in relation to suicidal ideation, anxiety, and mixed CMD outcomes was limited and inconsistent.

We found high-quality, consistent evidence that high baseline loneliness is associated with subsequent poorer depression outcomes among people with CMDs compared to those with low baseline loneliness. Most studies found that greater loneliness predicted more severe depressive symptoms, lower remission rates, or increased service use. This association was observed across clinical samples, community cohorts, and intervention trials, suggesting prognostic relevance in diverse contexts. Most studies were rated to be of high methodological quality, as they employed validated measures and adjusted for key confounders. Effect sizes were generally in the moderate magnitude range. While two studies reported null or negative findings, these appeared to reflect population-specific factors or intervention-related differences, rather than a contradiction of the overall pattern of effects.

In contrast, the evidence linking loneliness to suicidal ideation suggests that there is no consistent association between loneliness and future suicidal ideation in people with CMDs. Only one of five studies found a positive association, while the others reported null or negative results. Variability in study design, small sample sizes, and population characteristics likely contributed to this inconsistency. Findings for anxiety outcomes were limited and mixed. One study reported no association between baseline loneliness and subsequent anxiety symptoms after adjustment, while another found mixed effects for service utilisation in people with anxiety, with positive associations between baseline loneliness and face-to-face outpatient and community contacts at follow-up. The evidence for mixed CMD outcomes came from a single study and remains inconclusive.

With respect to treatment outcomes, only two studies evaluated whether loneliness affected intervention response. One study found that baseline loneliness was associated with poorer response to a peer-delivered CBT programme for women with physical disabilities and depression, while another found no association in a digital self-guided intervention for young adults with suicidal ideation. Further research is needed to clarify whether and how loneliness influences treatment response in CMDs.

Overall, these findings suggest that loneliness is associated with subsequent depression outcomes, however further work is needed to understand its role across CMDs and within treatment settings.

### Putting findings into context

Our findings align with theoretical models and systematic reviews that position loneliness as both a precursor and a perpetuating factor in the development and maintenance of depression [16, 17, 23]. The last review on this topic from Wang et al. [23] reviewed 34 longitudinal studies and found evidence that poor perceived social support and greater loneliness was associated with poorer depression outcomes. However, only two studies in that review assessed loneliness specifically. Our review expands these findings by providing a more focused synthesis, specifically on loneliness. It includes a larger number of relevant depression studies with longer follow-up durations, offering greater certainty about the longitudinal associations. A related review by Mann et al. [17] synthesised evidence from longitudinal studies investigating the relationship between loneliness and the onset of mental health problems in the general population, finding that loneliness at baseline is associated with the subsequent onset of depression. Our findings complement these by focusing on individuals already affected by CMDs, highlighting the role of loneliness in the persistence and severity of depression outcomes. Notably, our review found some evidence that loneliness may contribute to increased demand on mental health services for people with depression, suggesting that lonely individuals may continue to struggle despite engagement with care. Although preliminary, these findings raise important questions about the adequacy of standard treatments in addressing the needs of people with CMDs who are also lonely.

The consistent association between loneliness and poorer depression outcomes may reflect several underlying mechanisms. Loneliness is associated with increased rumination, negative social cognition, and reduced engagement in rewarding activities [58]. Ruminative thinking about being lonely has been found to be a mediator between loneliness and depressive symptoms [59]. Negative social cognition has also been proposed as a way through which loneliness might contribute to depressive symptoms via fostering feelings of rejection and social withdrawal [60]. Moreover, chronic loneliness has been linked to dysregulated stress responses, inflammation, and altered neural functioning, which may biologically predispose individuals to more severe or persistent depressive symptoms [61]. Recent evidence from a large Mendelian randomisation study found that, unlike many physical conditions, where observational associations with loneliness were not supported genetically, depression stood out as a condition where both lines of evidence align. This suggests that loneliness is not just a correlate but may be a causal contributor to depression [62]. In addition to these intra-personal processes, loneliness may reflect a lack of supportive others to encourage help-seeking or re-engagement with social and occupational roles. The absence of social companions may also reduce opportunities for meaningful activity, thereby undermining informal behavioural activation—an evidence-based mechanism for depression recovery [63]. These findings underscore the importance of identifying and addressing loneliness in individuals with depression. Interventions that target both the social and cognitive features of loneliness may be critical to improving long-term outcomes and should be prioritised in future research.

Our review did not find consistent evidence for a prospective association between loneliness and suicidal ideation in individuals with CMDs. This contrasts with prior reviews, such as McClelland et al. [12], which reported generally consistent positive associations between loneliness and suicidal ideation and behaviour across general and clinical populations, though with varying effect sizes and mediation by depression. In our review, which focused exclusively on longitudinal studies in clinical CMD samples, only one study reported a significant association between loneliness and suicidal ideation. This discrepancy may reflect differences in study design, populations, follow-up duration, and measurement strategies. It is also possible that loneliness influences suicidal ideation indirectly, through mediators such as hopelessness or depression, as proposed by the interpersonal theory of suicide and systematic review data [12, 64, 65].

Furthermore, a recent systematic review by Lu et al. [66] identified a moderate association between loneliness and suicidality in healthy adults and individuals with depressive disorders, based primarily on cross-sectional studies. However, our findings suggest that this relationship may not reliably extend to longitudinal associations in CMD populations. Unlike our review, which focused exclusively on longitudinal studies in clinical CMD samples, these broader reviews included mixed populations and—for Lu et al. [66] —cross-sectional designs. Our results should be interpreted with caution, as the GRADE certainty of evidence was low due to the limited number of longitudinal studies, inconsistent findings, and small sample sizes. These differences highlight the importance of standardised methodologies and longer-term studies to clarify the prospective role of loneliness in suicidal ideation among people with CMDs.

The mixed findings for anxiety outcomes in our review are consistent with the patterns observed in the broader literature, where weaker or less consistent associations between loneliness and anxiety are noted as compared to depression [23, 67, 68]. A cross-sectional study by Beutel et al. [67] found that loneliness was more strongly associated with depression than anxiety in general populations, potentially due to the distinct cognitive and social mechanisms underlying these disorders. The null finding reported by one study in our review, after adjustment for confounders, suggests that loneliness may interact with other factors, such as baseline anxiety severity or co-occurring depression, complicating its prognostic role.

### Strengths and limitations

Key strengths of this review include a comprehensive search strategy, inclusion of diverse CMD populations and settings, and detailed independent quality and certainty assessments using the NOS and GRADE criteria. We also screened and included studies published in languages other than English, reducing language bias and increasing the comprehensiveness of the review. Despite these strengths, there are a few limitations that should be acknowledged. First, our eligibility criteria—while intentionally broad in terms of outcome types and population characteristics—may have introduced heterogeneity that limited the comparability of studies and precluded meta-analysis. Second, few studies distinguished between types of loneliness (e.g., emotional and social), meaning we could not examine whether different dimensions of loneliness are differentially associated with outcomes. Third, in observational studies, the baseline often represents an arbitrary point in an individual’s illness trajectory. This makes it difficult to determine temporal precedence with certainty, particularly given the known bidirectionality between loneliness and CMD symptoms.

Nevertheless, longitudinal designs remain superior to cross-sectional studies in terms of causal inference, even if they do not capture full histories. Furthermore, heterogeneity in treatments received by participants, especially in non-intervention studies, may have confounded associations. In the two intervention studies, pooling experimental and control groups to assess the influence of loneliness may have diluted effects, as control groups often received no active treatment, potentially weakening observed associations. Finally, several included studies—many of those contributing to the evidence on depression—focused on older adult populations. This may limit the generalisability of findings to younger or more diverse populations with CMDs.

### Implications for Research

This systematic review highlights an important research gap: the absence of longitudinal evidence for the predictive role of loneliness in CMD outcomes beyond depression. Understanding whether loneliness worsens anxiety or other mental health outcomes over time is crucial for determining which clinical groups may benefit most from loneliness-targeted interventions.

Future research should also investigate how loneliness interacts with psychological therapies for CMDs. Most studies reviewed here focused on naturalistic outcomes, and very few examined whether loneliness moderates or mediates treatment response. Given the established links between loneliness and depression, we need to know how best to support depressed people who are also lonely. If loneliness is consistently associated with poorer treatment response - particularly in depression—this would support the development of bespoke interventions for people who are both lonely and depressed. Promising programmes, such as Community Navigators—a novel co-produced social intervention aiming to reduce loneliness in people with treatment resistant depression—show early potential and require further testing in full-scale RCTs [69, 70].

### Implications for Practice

Despite the current paucity of robust, evidence-based interventions specifically targeting loneliness in clinical populations, mental health professionals should not overlook its importance in clinical formulation and treatment planning.

The consistent association between loneliness and depression—both as a risk factor for its onset and as a prognostic factor for poorer outcomes—highlights the importance of routinely assessing loneliness as part of standard clinical care, particularly for individuals with depression. Even in the absence of formalised interventions, clinicians can adopt a proactive and curious stance by engaging patients in open conversations about loneliness and collaboratively identifying meaningful actions, such as community engagement, peer support, or social prescribing options. Given that loneliness is both common and modifiable, it warrants greater attention in treatment planning and offers an important opportunity to enhance mental health outcomes.

### Conclusions

This systematic review found consistent longitudinal evidence that loneliness is associated with subsequent poorer depression outcomes in individuals with CMDs. These associations were observed across clinical and non-clinical settings, and across diverse population groups, suggesting a robust and generalisable effect. In contrast, evidence for anxiety, suicidal ideation, and treatment outcomes was limited and mixed, highlighting important gaps in the current literature. Given the high prevalence of loneliness and its role in sustaining depression, there is a clear need for further research into interventions that address loneliness in people with CMDs. Targeting loneliness within mental health care may represent a promising strategy to improve clinical outcomes, particularly for depression, and should be prioritised in future research, policy, and practice.

## Data Availability

All data produced in the present work are contained in the manuscript and its supplementary information files.

https://www.crd.york.ac.uk/prospero/display_record.php?ID=CRD42023410401

## List of abbreviations

CMDs: Common mental disorders
NICE: National Institute for Health and Care Excellence
GAD: Generalised anxiety disorder
OCD: Obsessive-compulsive disorder
PTSD: Post-traumatic stress disorder
CORE: Centre for Outcomes and Research and Effectiveness
NOS: Newcastle-Ottawa Scale
RCTs: Randomised Controlled Trials
GRADE: Grading of Recommendations Assessment, Development and Evaluation
PHQ-9: Patient Health Questionnaire-9
CES-D-10: Center for Epidemiologic Studies Depression Scale
CIDI: Composite International Diagnostic Interview
NLP: Natural Language Processing
MDD: Major Depressive Disorder
NESDO: The Netherlands Study of Depression in Older Persons
NESDA: The Netherlands Study of Depression and Anxiety
DBT: Dialectical Behaviour Therapy
ACT: Acceptance and Commitment Therapy
CBT: Cognitive behavioural therapy
BDI-II: Beck Depression Inventory

## Declarations

### Ethics approval and consent to participate

Not applicable. This study is a systematic review of previously published research and does not involve human participants or require ethical approval.

### Consent for publication

Not applicable.

### Availability of data and materials

All data generated or analysed during this study are included in this published article and its supplementary information files.

### Authors’ contributions

TS conceived the study with BLE and SJ. TS led the protocol development, database search, screening, data extraction, analysis, quality appraisal, certainty of evidence assessments, and manuscript drafting. BLE, SJ, AP, GL and JB provided methodological guidance and supervised the review. TS, GE, MO, ST, JHS, HG and KL conducted screening. TS, HG and KL conducted data extraction. TS and HG performed the quality appraisal assessments. TS conducted the narrative synthesis and certainty of evidence assessments with input from BLE and SJ. TS drafted the manuscript. All authors reviewed and approved the final manuscript.

## Acknowledgements

We thank Phoebe Barnett from the UCL CORE Unit for her valuable advice during the review process. We are also grateful to Antonio Rojas García and Maria Ana Matias for assisting with the translation of non-English articles, and to Miriam Nyawira for her help with title and abstract screening.

