## Appendices for "Associations between loneliness and outcomes of Common Mental Disorders (CMDs): A systematic review of longitudinal studies"

### Appendix 1: Prisma Checklist

| Section and Topic | Item # | Checklist item | Location where item is reported |
|---|---|---|---|
| **TITLE** | | | |
| Title | 1 | Identify the report as a systematic review. | Page 1 |
| **ABSTRACT** | | | |
| Abstract | 2 | See the PRISMA 2020 for Abstracts checklist. | Page 2 |
| **INTRODUCTION** | | | |
| Rationale | 3 | Describe the rationale for the review in the context of existing knowledge. | Pages 3-5 |
| Objectives | 4 | Provide an explicit statement of the objective(s) or question(s) the review addresses. | Pages 6 |
| **METHODS** | | | |
| Eligibility criteria | 5 | Specify the inclusion and exclusion criteria for the review and how studies were grouped for the syntheses. | Pages 7-9 |
| Information sources | 6 | Specify all databases, registers, websites, organisations, reference lists and other sources searched or consulted to identify studies. Specify the date when each source was last searched or consulted. | Page 9 |
| Search strategy | 7 | Present the full search strategies for all databases, registers and websites, including any filters and limits used. | Page 9 and Appendix |
| Selection process | 8 | Specify the methods used to decide whether a study met the inclusion criteria of the review, including how many reviewers screened each record and each report retrieved, whether they worked independently, and if applicable, details of automation tools used in the process. | Page 10 |
| Data collection process | 9 | Specify the methods used to collect data from reports, including how many reviewers collected data from each report, whether they worked independently, any processes for obtaining or confirming data from study investigators, and if applicable, details of automation tools used in the process. | Page 10 |
| Data items | 10a | List and define all outcomes for which data were sought. Specify whether all results that were compatible with each outcome domain in each study were sought (e.g. for all measures, time points, analyses), and if not, the methods used to decide which results to collect. | Page 12 |
| | 10b | List and define all other variables for which data were sought (e.g. participant and intervention characteristics, funding sources). Describe any assumptions made about any missing or unclear information. | Page 12 |
| Study risk of bias assessment | 11 | Specify the methods used to assess risk of bias in the included studies, including details of the tool(s) used, how many reviewers assessed each study and whether they worked independently, and if applicable, details of automation tools used in the process. | Page 11 |
| Effect measures | 12 | Specify for each outcome the effect measure(s) (e.g. risk ratio, mean difference) used in the synthesis or presentation of results. | Page 12 |
| Synthesis methods | 13a | Describe the processes used to decide which studies were eligible for each synthesis (e.g. tabulating the study intervention characteristics and comparing against the planned groups for each synthesis (item #5)). | Pages 11-12 |
| | 13b | Describe any methods required to prepare the data for presentation or synthesis, such as handling of missing summary statistics, or data conversions. | Pages 11-12 |
| | 13c | Describe any methods used to tabulate or visually display results of individual studies and syntheses. | Pages 11-12 |
| | 13d | Describe any methods used to synthesize results and provide a rationale for the choice(s). If meta-analysis was performed, describe the model(s), method(s) to identify the presence and extent of statistical heterogeneity, and software package(s) used. | Pages 11-12 |
| | 13e | Describe any methods used to explore possible causes of heterogeneity among study results (e.g. subgroup analysis, meta-regression). | Pages 11-12 |

| | | | |
|---|---|---|---|
| | 13f | Describe any sensitivity analyses conducted to assess robustness of the synthesized results. | Pages 11-12 |
| Reporting bias assessment | 14 | Describe any methods used to assess risk of bias due to missing results in a synthesis (arising from reporting biases). | Page 11 |
| Certainty assessment | 15 | Describe any methods used to assess certainty (or confidence) in the body of evidence for an outcome. | Page 11 |
| **RESULTS** | | | |
| Study selection | 16a | Describe the results of the search and selection process, from the number of records identified in the search to the number of studies included in the review, ideally using a flow diagram. | Page 13 and Figure 1 |
| | 16b | Cite studies that might appear to meet the inclusion criteria, but which were excluded, and explain why they were excluded. | Page 14 and Appendix |
| Study characteristics | 17 | Cite each included study and present its characteristics. | Page 14 and Table 1 |
| Risk of bias in studies | 18 | Present assessments of risk of bias for each included study. | Page 15 and Appendix |
| Results of individual studies | 19 | For all outcomes, present, for each study: (a) summary statistics for each group (where appropriate) and (b) an effect estimate and its precision (e.g. confidence/credible interval), ideally using structured tables or plots. | Table 1, Table 2, Table 3, Table 4 |
| Results of syntheses | 20a | For each synthesis, briefly summarise the characteristics and risk of bias among contributing studies. | Pages 14 - 21 |
| | 20b | Present results of all statistical syntheses conducted. If meta-analysis was done, present for each the summary estimate and its precision (e.g. confidence/credible interval) and measures of statistical heterogeneity. If comparing groups, describe the direction of the effect. | Pages 14 - 21 |
| | 20c | Present results of all investigations of possible causes of heterogeneity among study results. | Pages 14 - 21 |
| | 20d | Present results of all sensitivity analyses conducted to assess the robustness of the synthesized results. | Pages 14 - 21 |
| Reporting biases | 21 | Present assessments of risk of bias due to missing results (arising from reporting biases) for each synthesis assessed. | |
| Certainty of evidence | 22 | Present assessments of certainty (or confidence) in the body of evidence for each outcome assessed. | Pages 14 - 21 |
| **DISCUSSION** | | | |
| Discussion | 23a | Provide a general interpretation of the results in the context of other evidence. | Page 23-25 |
| | 23b | Discuss any limitations of the evidence included in the review. | Pages 25-26 |
| | 23c | Discuss any limitations of the review processes used. | Pages 25-26 |
| | 23d | Discuss implications of the results for practice, policy, and future research. | Pages 26-28 |
| **OTHER INFORMATION** | | | |
| Registration and protocol | 24a | Provide registration information for the review, including register name and registration number, or state that the review was not registered. | Page 7 |
| | 24b | Indicate where the review protocol can be accessed, or state that a protocol was not prepared. | Page 7 |
| | 24c | Describe and explain any amendments to information provided at registration or in the protocol. | Not applicable |

| | | | |
|---|---|---|---|
| Support | 25 | Describe sources of financial or non-financial support for the review, and the role of the funders or sponsors in the review. | Page 30 |
| Competing interests | 26 | Declare any competing interests of review authors. | Page 29 |
| Availability of data, code and other materials | 27 | Report which of the following are publicly available and where they can be found: template data collection forms; data extracted from included studies; data used for all analyses; analytic code; any other materials used in the review. | Page 29 |

**Appendix 2: Full search strategy**

| Database | Records retrieved 2023 | Updated search 2024 |
|---|---|---|
| MEDLINE via Ovid | 2316 | 242 (23032023 – 13052024) |
| PsycINFO via Ovid | 1652 | 305 (08032023 – 13052024) |
| Embase via Ovid | 2159 | 568 (11032023 – 13052024) |
| CINAHL via EBSCOhost | 1540 | 304 (11032023 – 13052024) |
| TOTAL | 7667 | 1419 |
| Duplicates | 1727 | 249 |
| Records to be screened | 5940 | 1170 |

**Ovid MEDLINE(R) ALL <1946 to March 22, 2023>**

1 Loneliness/ 6053

2 ((loneliness or lonel* or lonely or social*) adj4 isolat*).mp. or confiding relationship*.ti,ab,kf. [mp=title, book title, abstract, original title, name of substance word, subject heading word, floating sub-heading word, keyword heading word, organism supplementary concept word, protocol supplementary concept word, rare disease supplementary concept word, unique identifier, synonyms, population supplementary concept word, anatomy supplementary concept word] 26928

3 ("common mental disorder*" or CMD or "mental disorder*" or mental or psychiatr* or depression or anxiety or depress* or anxious or anxiet* or "anxiety disorder*" or phobia* or OCD or "obsessive-compulsive disorder*" or PTSD or "post-traumatic stress disorder*" or "panic disorder*").mp. or panic attack*.ti,ab,kf. [mp=title, book title, abstract, original title, name of substance word, subject heading word, floating sub-heading word, keyword heading word, organism supplementary concept word, protocol supplementary concept word, rare disease supplementary concept word, unique identifier, synonyms, population supplementary concept word, anatomy supplementary concept word] 1520993

4 Mental Disorders/ 176520

5 anxiety disorders/ or anxiety, separation/ or neurocirculatory asthenia/ or neurotic disorders/ or obsessive-compulsive disorder/ or phobic disorders/ 82838

6 Depression/ 148252

7 3 or 5 or 6 1533475

8 Longitudinal Studies/ 163821

9 Case-Control Studies/ 326749

10 Retrospective Studies/ 1103411

11 Prospective Studies/ 653991

12 (cohort or longitudinal or prospective or retrospective or registr* or "prospective stud*" or "retrospective stud*" or "case* adj5 control*" or "case adj3 comparison*" or "case-comparison" or "control group*" or RCT or trial or "intervention stud*" or "randomi#ed controlled trial*" or "comparison stud*" or "non-randomi#ed trial" or "quasi-experimental trial*").mp. or "pre-post stud*".ti,ab,kf. [mp=title, book title, abstract, original title, name of substance word, subject heading word, floating sub-heading word, keyword heading word, organism supplementary concept word, protocol supplementary concept word, rare disease supplementary concept word, unique identifier, synonyms, population supplementary concept word, anatomy supplementary concept word] 4615206

13 8 or 9 or 10 or 11 or 12 4804280

14 1 or 2 31288

15 7 and 13 and 14 2662

16 limit 15 to humans 2316

17 limit 16 to dt=20230323-20240513 242

**APA PsycInfo <1806 to March Week 2 2023>**

1 exp Loneliness/ 6370

2 (loneliness or lonel* or lonely or "social* adj4 isolat*").mp. or "confiding relationship*".ti,ab,id. [mp=title, abstract, heading word, table of contents, key concepts, original title, tests & measures, mesh word] 16309

3 1 or 2 16309

4 ("common mental disorder*" or CMD or "mental disorder*" or mental or psychiatr* or depression or anxiety or depress* or anxious or anxiet* or "anxiety disorder*" or phobia* or OCD or "obsessive-compulsive disorder*" or PTSD or "post-traumatic stress disorder*" or "panic disorder*").mp. or "panic attack*".ti,ab,id. [mp=title, abstract, heading word, table of contents, key concepts, original title, tests & measures, mesh word] 1258760

5 exp Mental Disorders/ 1013486

6 anxiety disorders/ or mental disorders/ or castration anxiety/ or generalized anxiety disorder/ or panic attack/ or panic disorder/ or phobias/ or separation anxiety disorder/ or anxiety management/ or anxiety screening/ or obsessive compulsive disorder/ or social anxiety/ 149051

7 exp Late Life Depression/ or exp Recurrent Depression/ or exp "Depression (Emotion)"/ or exp "Long-term Depression (Neuronal)"/ or exp Postpartum Depression/ or exp Treatment Resistant Depression/ or exp Major Depression/ or exp Beck Depression Inventory/ or exp Depression Screening/ or exp Atypical Depression/ or exp Reactive Depression/ 182417

8 4 or 5 or 6 or 7 1702319

9 exp Longitudinal Studies/ 17354

10 exp Prospective Studies/ 1296

11 exp Retrospective Studies/ 910

12 (cohort or longitudinal or prospective or retrospective or registr* or "prospective stud*" or "retrospective stud*" or "case* adj5 control*" or "case adj3 comparison*" or "case-comparison" or "control group*" or RCT or trial or "intervention stud*" or "randomi#ed controlled trial*" or

"comparison stud*" or "non-randomi#ed trial*" or "quasi-experiment*").mp. or "pre-post stud*".ti,ab,id. [mp=title, abstract, heading word, table of contents, key concepts, original title, tests & measures, mesh word] 569755

13 9 or 10 or 11 or 12 569755

14 3 and 8 and 13 1791

15 limit 14 to human 1652

16 limit 15 to up=20230308-20240513 305

**Embase <1980 to 2023 Week 11>**

1 loneliness/ or de Jong Gierveld Loneliness Scale/ or UCLA Loneliness Scale/ or Revised UCLA Loneliness Scale/ 13802

2 (loneliness or lonel* or lonely or "social* adj4 isolat*").mp. or "confiding relationship*".ti,ab,kf. [mp=title, abstract, heading word, drug trade name, original title, device manufacturer, drug manufacturer, device trade name, keyword heading word, floating subheading word, candidate term word] 18245

3 1 or 2 18245

4 ("common mental disorder*" or CMD or "mental disorder*" or mental or psychiatr* or depression or anxiety or depress* or anxious or anxiety* disorder* or phobia* or OCD or "obsessive compulsive disorder*" or PTSD or "post-traumatic stress disorder" or "panic disorder*").mp. or "panic attack*".ti,ab,kf. [mp=title, abstract, heading word, drug trade name, original title, device manufacturer, drug manufacturer, device trade name, keyword heading word, floating subheading word, candidate term word] 2076254

5 depression/ or anxiety disorder/ or mental disease/ 709721

6 4 or 5 2076254

7 (cohort adj (study or studies)).tw. 456601

8 (case control adj (study or studies)).tw. 166514

9 (observational adj (study or studies)).tw.  248098

10 (epidemiologic* adj (study or studies)).tw.  119073

11 Case control study/  203349

12 longitudinal study/  189579

13 retrospective study/ 1433158

14 prospective study/ 859032

15 randomized controlled trials/ 255142

16 (RCT or trial or "intervention stud*" or "comparison stud*" or "non-randomi#ed trial" or "quasi-experimental trial").mp. or "pre-post stud*".tw. [mp=title, abstract, heading word, drug trade name, original title, device manufacturer, drug manufacturer, device trade name, keyword heading word, floating subheading word, candidate term word]  2622149

17 7 or 8 or 9 or 10 or 11 or 12 or 13 or 14 or 15 or 16.   5243607

18   3 and 6 and 17   2175

19   limit 18 to human 2159

20 limit 19 to dc=20230311-20240513   568

**CINAHL search ran on 22/03/2023 (Limiters: Exclude MEDLINE records; Human)**

S1 MH loneliness 6,481

S2 ((TI "Mood disorder*" OR AB "Mood disorder*" OR SU "Mood disorder*") OR (TI cyclothymi* OR AB cyclothymi* OR SU cyclothymi*) OR (TI depression OR AB depression OR SU depression) OR (TI depressive OR AB depressive OR SU depressive) OR (TI dysthymi* OR AB dysthymi* OR SU dysthymi*) OR (TI neurotic OR AB neurotic OR SU neurotic) OR (TI neurosis OR AB neurosis OR SU neurosis) OR (TI "adjustment disorder*" OR AB "adjustment disorder*" OR SU "adjustment disorder*") OR (TI "anxiety disorder*" OR AB "anxiety disorder*" OR SU "anxiety disorder*") OR (TI anxious OR AB anxious OR SU anxious) OR (TI "health anxiety" OR AB "health anxiety" OR SU "health anxiety") OR (TI agoraphobia OR AB agoraphobia OR SU agoraphobia) OR (TI obsess* OR AB obsess* OR SU obsess*) OR (TI compulsi* OR AB compulsi* OR SU compulsi*) OR (TI OCD OR AB OCD OR SU OCD) OR (TI OCPD OR AB OCPD OR SU OCPD) OR (TI panic OR AB panic OR SU panic) OR (TI phobi* OR AB phobi* OR SU phobi*) OR (TI ptsd OR AB ptsd OR SU ptsd) OR (TI posttrauma* OR AB posttrauma* OR SU posttrauma*) OR (TI "post trauma*" OR AB "post trauma*" OR SU "post trauma*") OR (TI "body dysmorphi*" OR AB "body dysmorphi*" OR SU "body dysmorphi*") OR (TI "affective symptoms" OR AB "affective symptoms" OR SU "affective symptoms") OR (((TI mental* OR AB mental* OR SU mental*) OR (TI psychologic* OR AB psychologic* OR SU psychologic*)) W1 ((TI health OR AB health OR SU health) OR (TI well* OR AB well* OR SU well*)))) 455,081

S3 (MH "Depression") OR (MH "Dysthymic Disorder") OR (MH "Anxiety Disorders") OR (MH "Generalized Anxiety Disorder") OR (MH "Obsessive-Compulsive Disorder") OR (MH "Panic Disorder") OR (MH "Agoraphobia") OR (MH "Phobic Disorders") OR (MH "Social Anxiety Disorders") OR (MH "Stress Disorders, Post-Traumatic") 165,730

S4 (((AB loneliness OR TI loneliness OR SU loneliness) OR (AB lonel* OR TI lonel* OR SU lonel*) OR (AB lonely OR TI lonely OR SU lonely) OR ((AB social* OR TI social* OR SU social*) N4 (AB isolate* OR TI isolat* OR SU isolate*)) OR (AB "confiding relationship*" OR TI "confiding relationship*" OR SU "confiding relationship*"))) 11,450

S5 S1 OR S4 11,450

S6 S2 OR S3 455,081

S7 S5 AND S6 4,727

S8 MH cohort study 0

S9 MH longitudinal 0

S10 (MH "Epidemiological Research") OR (MH "Prospective Studies") OR (MH "Cross Sectional Studies") OR (MH "Case Control Studies") OR (MH "Retrospective Design") OR (MH "Repeated Measures")

S11 (TI "Case control" OR AB "Case control") 42,510

S12 ((TI cohort OR AB cohort) W1 ((TI study OR AB study) OR (TI studies OR AB studies))) 126,788

S13 (TI Longitudinal OR AB Longitudinal) 112,409

S14 (MH "Clinical Trials+") 349,746

S15 TX randomi* control* trial* 260,342

S16 TX non randomi* control* trial* 3,046

S17 TX quasi experim* 21,773

S18 S8 OR S9 OR S10 OR S12 OR S13 OR S14 OR S15 OR S16 OR S17 1,523,182

S19 S5 AND S6 AND S18 1,540

S20 EM '20230311- 20240513' 294,416

S21 S19 AND S39 304

**Appendix 3: Screening guides**

**A. Title and abstract screening guide for Loneliness Systematic Review**

1. Before starting any screening, please make sure you read through the protocol carefully. Here is a link to it: https://www.crd.york.ac.uk/prospero/display_record.php?ID=CRD42023410401

2. At this stage, we are only screening titles and abstracts – you should not view any full-text articles at this point.

3. If you are not sure about a paper for this stage, please mark it as included (NOT maybe). This way we can check it more thoroughly at the full-text stage.

4. Please screen independently – it's important not to discuss any paper with someone else on the screening team.

5. When excluding a paper, please assign it only one exclusion reason. If there is more than one reason that you think the paper doesn't fit, please select the reason that is highest in this order:
    1. Duplicate
    2. Not a research study (e.g., book chapter, conference paper, editorial)
    3. Wrong study type (Cross-sectional studies, qualitative studies, systematic reviews and meta-analyses)
    4. Wrong population (Children under 16 years old, or populations with organic disorders; studies concerning people with drug and alcohol disorders only and no other major psychiatric disorder, or populations with primary diagnosis of severe mental illness (SMI) (Psychosis, bipolar disorder, depression with psychotic features, personality disorders), or CMDs secondary to or following the onset of neurological or neurodegenerative disorders/diseases (e.g. traumatic brain injuries, stroke, dementias, Parkinson's disease)
    5. Studies evaluating social isolation and related concepts
    6. Mental health outcomes not the primary outcome
    7. Relevant review

6. Screening questions:

These are sorted in order of importance so that a reference can be excluded as soon as it fails to meet a criterion

- Is it a research study?
- Was the study longitudinal? (Both observational studies and intervention studies)
- Was some or all of the study population adults 16+ with CMD*?
- Was the study evaluating loneliness?

*According to NICE clinical guidance 2011 CMDs include depression and anxiety disorders such as generalised anxiety disorder, panic disorder, mixed depression and anxiety, obsessive-compulsive disorder (OCD) and post-traumatic stress disorder (PTSD) as well as phobias about specific things or situations.

### B. Full-text screening Guide for Loneliness Systematic Review

Introduction: This guide is designed to assist reviewers in the full-text screening process using Rayyan. Please refer to these criteria when evaluating each study. Remember to apply these criteria consistently and to flag any uncertainties for discussion with the review team.

How to Use This Guide:

1. Log into Rayyan and access the assigned studies.
2. For each study, review the full text with these criteria in mind.
3. Use Rayyan's system to mark studies as Include, Exclude, or Maybe.
4. Use Rayyan's notes feature to briefly explain exclusions or flag uncertainties.

Inclusion Criteria: Studies must meet all of the following criteria to be included:

1. Study Design: Longitudinal study (observational or intervention) □ Dissertations (considered as grey literature) are eligible if they meet all other criteria
2. Population: Adults aged 16 or older □ Participants have common mental disorders (CMDs)

- For general population samples:
    - At least 50% of participants should meet CMD symptom threshold
    - Use clinical cut-offs specific to each screening tool/measure
    - Include Studies with Subgroup Analysis – If a study reports findings separately for those meeting CMD criteria, it could be included, even if the total sample doesn't meet 50%.
- For clinical samples:
    - Participants should meet diagnostic criteria or clinical cut-offs for CMDs

3. Exposure: Measures loneliness at baseline using a recognized scale
4. Outcomes: Reports at least one of the following at follow-up:
    - CMD symptom severity (including suicidality)
    - Service use (hospital/crisis admission/CMHTs)
    - Quality of life
    - Social functioning
    - Personal recovery
    - Relapse after remission
    - Wellbeing
    - Global mental health
5. Analysis: Reports quantitative association between baseline loneliness (predictor) and at least one of the outcomes listed above at follow-up □ For general population studies, analysis must focus on the sub-sample meeting CMD criteria at baseline □ Mediation studies are eligible if they meet other criteria (flag these for separate analysis)
6. Language: Full text available in English or a European language that can be translated
7. Special Considerations: COVID-19 studies are eligible if they meet other criteria (flag these for separate analysis)

Exclusion Criteria: Exclude studies that meet ANY of the following criteria:

□ Cross-sectional design □ Qualitative study □ Systematic review or meta-analysis □ Focus exclusively on people with drug and alcohol disorders □ Focus primarily on severe mental illness (SMI) □ Focus on CMDs secondary to neurological or neurodegenerative disorders □ Includes children under 16 years old □ Does not analyse loneliness as a predictor of CMD outcomes

Decision Making:

- Include: Study meets all inclusion criteria and no exclusion criteria

- Exclude: Study fails to meet any inclusion criterion or meets any exclusion criterion
- Maybe: Uncertain about any criterion; flag for team discussion

Exclusion Process: When excluding a paper, assign only one exclusion reason. If there are multiple reasons for exclusion, select the reason that is highest in the following order:

1. Duplicate
2. Wrong publication type (not a research study)
3. Wrong study design
4. Wrong population
5. Study did not evaluate loneliness
6. Wrong outcome
7. Relevant review
8. Incorrect Association (If the study does not address the correct relationship between loneliness and mental disorders as defined in your review criteria.)

Handling Uncertainties:

- Use Rayyan's notes feature to explain your uncertainty
- Tag the study for discussion in Rayyan
- Consult with a second reviewer or the project lead for resolution

Remember:

- Be thorough but efficient
- Consistency is key, especially in assigning exclusion reasons
- When in doubt, mark as 'Maybe' and explain your reasoning
- Flag mediation studies and COVID-19 studies for separate analysis in the write-up

For any questions about this process, please contact Dora Stefanidou;

**Appendix 4: NOS**

### Adapted Newcastle–Ottawa Scale (NOS) for Loneliness Review

### COHORT STUDIES

Note: We modified the Cohort Studies scale to fit the focus of this review. The population of interest in this review is adults 16+ with any CMD and the exposure is loneliness.

Scoring: A study can be awarded a maximum of one star for each numbered item within the Selection and Outcome categories. A maximum of two stars can be given for Comparability. Each study's score was calculated by summing the total of these scores. Based on total NOS scores, studies were rated as high quality (7–9 stars), moderate quality (5–6 stars), or low quality (0–4 stars).

Selection (Max 1 star per item)

1) Representativeness of the exposed cohort
   a) truly representative of the average individual with CMD in the community *
   b) somewhat representative of the average individual with CMD in the community *
   c) selected group of users (e.g. nurses, university students, veterans)
   d) no description of the derivation of the cohort

2) Selection of the non exposed cohort (adapted for loneliness as exposure)
   a) CMD participants compared across levels of loneliness from same population *
   b) drawn from a different source
   c) no description of the derivation of the non exposed cohort

3) Ascertainment of exposure
   a) secure record (e.g. medical records) or loneliness validated measure (e.g. UCLA, De Jong) *
   b) structured interview using validated tool*
   c) written self-report (single question)
   d) no description

4) Demonstration that outcome of interest was not present at start of study
   a) yes *
   b) no

Comparability (Max 2 stars)

1) Comparability of cohorts on the basis of the design or analysis
   a) study controls for age, sex, baseline mental health status *
   b) study controls for any additional factor *
   c) Cohorts are not comparable on the basis of the design or analysis controlled for confounders

Outcome (Max 1 star per item)

1) Assessment of outcome
   a) independent blind assessment *
   b) record linkage *
   c) self-report
   d) no description

2) Was follow-up long enough for outcomes to occur
   a) yes (≥ 3 months for mental health outcomes) *
   b) no

3) Adequacy of follow up of cohorts
   a) complete follow up - all subjects accounted for *
   b) subjects lost to follow up unlikely to introduce bias - small number lost - (LESS than or equal 30% or description provided of those lost) *
   c) follow up rate less than 70% and no description of those lost
   d) no statemen

**Reviewer Notes and Manual**

- Selection item 2: A star is awarded if the study includes a comparison group of individuals with CMDs who differ in loneliness levels (e.g., high vs low loneliness), and both groups are drawn from the same population. No star is given for studies without such a comparison group. This adaptation allows us to differentiate studies that can more robustly test the effect of loneliness (by comparing lonely vs non-lonely individuals with CMDs) from those that simply measure loneliness as a continuous predictor without group comparison.

- Selection item 3: Give a star if loneliness was measured using a validated tool such as the UCLA or De Jong Gierveld Loneliness Scales. No star is given if loneliness was assessed using a non-validated, vague, single-question measure, or unclear description of how it was measured.
- Selection item 4: Baseline CMD is always present – give a star always.
- Outcome item 1: A star is awarded if CMD outcomes were assessed using a validated measure (e.g., PHQ-9, GAD-7, MINI) or a structured clinical interview.

**Appendix 4: NOS Ratings**

| Study ID | Selection | Comparability | Outcome | Total | Quality |
|---|---|---|---|---|---|
| Antonelli-Salgado et al., 2021 | *** | ** | * | 6 | Moderate |
| Brown et al., 2023 | *** | * | * | 5 | Moderate |
| Chen et al., 2020 | ** | ** | *** | 7 | High |
| Gabarrell-Pascuet et al., 2022 | *** | ** | ** | 7 | High |
| Hassouneh et al., 2013 | ** | ** | *** | 7 | High |
| Holvast et al., 2015 | **** | ** | *** | 9 | High |
| Jeuring et al., 2018 | *** | ** | ** | 7 | High |
| Kivelä et al., 2019 | *** | ** | ** | 7 | High |
| Maarsingh et al., 2018 | *** | ** | *** | 8 | High |
| MacNeil et al., 2023 | ** | * | ** | 5 | Moderate |
| McGillivray et al., 2023 | *** | * | ** | 6 | Moderate |
| Nuyen et al., 2020 | **** | ** | ** | 8 | High |
| Ortiz et al., 2017 | *** | * | *** | 7 | High |
| Parmar et al., 2024 | **** | ** | *** | 9 | High |
| Schaakxs et al., 2018 | *** | ** | *** | 8 | High |
| van Beljouw et al., 2010 | *** | ** | *** | 8 | High |
| Van de Brink et al. 2018 | *** | ** | *** | 8 | High |

**Moderate n=4**
**High n=13**
**Sum n=17**

### Appendix 5: GRADE Certainty of evidence for each outcome

| No. of studies | Study quality | Concerns about certainty (serious concerns: >50% of information is from studies at low risk of bias) | Indirectness | Concerns about certainty | Imprecision | Concerns about certainty | Inconsistency | Concern about certainty | Publication bias | Concerns about certainty | Certainty |
|---|---|---|---|---|---|---|---|---|---|---|---|
| **Depression** | | | | | | | | | | | |
| 13 | 11/13 studies were of high methodological quality | No concerns | All the studies reported direct evidence - all studies included measures of loneliness and mental health outcomes in samples with CMDs. Most samples included clinical populations with CMDs, but a few | No concerns | Four studies had sample sizes below 400 (4/13 < 400) and nine studies had sample sizes above 400 (9/13 > 400). However, the studies with small sample sizes reported the same trends as studies with larger sample sizes except for one study. | Borderline | 85% (11/13) of studies consistently show loneliness is associated with worse depression outcomes (symptom severity, recurrence, or remission). Parmar et al.'s service utilization outcomes align with worse | No concerns | We do not suspect publication bias as both significant and non-significant findings were published | No concerns | High certainty ⊕⊕⊕⊕ |

| | | | | | | | | | | | |
|---|---|---|---|---|---|---|---|---|---|---|---|
| | | | were general population samples with more that 50% of participants experiencing CMDs. | | | | outcomes in terms of increased service demand, supporting the trend, though not symptom specific. Chen et al.'s contrary finding and McGillivray et al.'s null result reduce consistency. | | | | |
| **Suicidal ideation** | | | | | | | | | | | |
| 5 | 3/5 studies were of moderate quality, 2/5 were high quality | Borderline | The studies all reported direct evidence- all studies included measures of loneliness and mental health outcomes in samples with CMDs. Most samples included clinical | No concerns | Three studies had sample sizes below 400 (3/5 < 400) and two studies had sample sizes above 400 (2/5 > 400). Studies with small sample sizes reported no associations or reverse association. | Serious | 60% of the available evidence suggested no association between baseline loneliness and suicidal ideation at follow-up, with another 40% reporting varying | Serious | We do not suspect publication bias as both significant and non-significant findings were published. | No concerns | Low certainty ⊕⊕OO |

| | | | | | | | | | | | |
|---|---|---|---|---|---|---|---|---|---|---|---|
| | | | populations with CMDs, but a few were general population samples with more that 50% of participants experiencing CMDs. | | | | direction of effects. | | | | |
| **Anxiety** | | | | | | | | | | | |
| 2 | 2/2 studies were of high methodological quality | No concerns | The studies all reported direct evidence- all studies included measures of loneliness and mental health outcomes in samples with CMDs. Most samples included clinical populations with CMDs, but a few were general population samples with | No concerns | Only two studies. Both studies had a sample size above 400 (2/2 > 400) | Serious | Studies reported different directions of effect. One study reported a null effect and another one mixed effects (significant positive effect for contacts, null for emergency outcomes). Variability amplified by outcome type differences | Serious | Possible publication bias due to limited studies - only two | Serious | Very low certainty ⨁OOO |

| | | | | | | | | | | | |
|---|---|---|---|---|---|---|---|---|---|---|---|
| | | | more that 50% of participants experiencing CMDs. | | | | (service use vs. symptom severity). | | | | |
| **Mixed CMD outcomes** | | | | | | | | | | | |
| 1 | The single study was of high methodological quality | No concerns | N/A | N/A | Single study (below threshold). The study had a subgroup of participants with CMDs at baseline for whom a separate analysis was conducted. Sample size reported for the overall sample not for the subgroup. Loneliness and mental health outcomes were assessed using validated measures. | Serious | Differing findings in adjusted and unadjusted models with wide confidence intervals. | Serious | Possible publication bias due to limited studies - only one | Serious | Very low certainty ⨁OOO |

**Appendix 6: Near Misses**

Six studies were initially considered for inclusion in the review as they examined associations between loneliness and CMD outcomes (Jesus et al., 2024; Keller et al., 2023, O'Day et al., 2021, Southward et al., 2022, Walker at al., 2020, Wielaard at al., 2017). However, during the data extraction phase, it became evident that these associations were cross-sectional, not longitudinal, and thus did not meet the inclusion criteria. Two of these studies focused on mixed CMD outcomes, one on anxiety and three on depression. Of these, one general population study (Keller et al., 2023) found that loneliness and anxiety mediated the relationship between psychological distress and depressive symptoms during the COVID-19 pandemic, suggesting loneliness as a sustaining factor. Among studies involving mental health service users, Wielaard et al. (2017) found that loneliness significantly mediated the link between childhood abuse and depression diagnosis in older adults. Four intervention studies also offered relevant findings: Jesus et al. (2024) reported that a home-based psychotherapeutic program reduced depressive symptoms via reductions in social loneliness, Walker et al. (2020) assessed a Stepped Collaborative Care aiming to improve depression management in primary care settings and found that higher baseline loneliness was associated with depression severity at baseline; Southward et al. (2022) found that within-person increases in loneliness were associated with concurrent increases in anxiety and depression during therapy; and O'Day et al (2021) found that reductions in loneliness from pre- to post-treatment were significantly associated with lower levels of social anxiety during the follow-up period (3 to 12 months) in participants receiving cognitive-behavioural group therapy (CBGT) and mindfulness-based stress reduction (MBSR) to treat social anxiety disorders. A summary of the characteristics and findings of these studies can be found in Appendix 6 Table1 and Table 2.

**Table 1. Study Characteristics of Near Misses N=6**

| Author(s), Year | Country | Study Design | Sample size N | Population | CMD | CMD determined by | Age range Mean and (SD) | Gender N (%) | Ethnicity N (%) | Loneliness Measure | Follow-up duration |
|---|---|---|---|---|---|---|---|---|---|---|---|
| Jesus et al., 2024 | Portugal | RCT analysed as prospective cohort study | 199 (HEPPI: 98, TAU: 101) | Homebound older adults presenting mild cognitive impairment and psychological symptomatology | Depression and anxiety symptoms | ≥ 11 on the Geriatric Depression Scale-30 (GDS-30); ≥ 8 on the Geriatric Anxiety Inventory (GAI) | R: 65+ years<br><br>HEPPI group: 79.12 (5.44); TAU group: 80.27 (4.68) | Female N=165 (83.01%)<br>Male N=34 (16.99%) | NR | UCLA Loneliness Scale (Portuguese version) | 3 months post-intervention |
| Keller et al., 2023 | Germany | Longitudinal cohort study | 403 | Psychosomatic rehabilitation patients | Depression and anxiety symptoms | PHQ-2, GAD-2 | R: 18–60+ years<br><br>39 years or younger:12.2 %<br><br>Between 40 and 49: 20.9%<br><br>Between 50 and 59 years of age: 51%<br><br>60 years or older: 15.9% | Female N=264 (65.8%) | NR | One item from the UCLA Loneliness Scale and the Loneliness Item from CES-D | 6 weeks pre-rehabilitation to 12 weeks post-rehabilitation |

| | | | | | | | | | | | |
|---|---|---|---|---|---|---|---|---|---|---|---|
| O'Day et al., 2021 | USA | RCT (analysed as a prospective cohort study) | 108 | Treatment-seeking individuals with social anxiety disorder | Social anxiety disorder | Anxiety Disorders Interview Schedule for the DSM-IV: Lifetime version (ADIS-IV-L) | 32.7 (8.0) | Female: 55.6 % | Caucasian/ White: 47 (43.5%) Asian American: 42 (38.9%) Latino/Hispanic: 10 (9.3%) African American/ Black: 1 (0.9%) American Indian/Alaskan Native: 1 (0.9%) More than one race: 7 (6.5%) | UCLA-8 | Post-treatment, 3, 6, 9 and 12 months |
| Southward et al. 2022 | USA | Secondary data analysis of a SMART trial | 70 | Clinical population with anxiety, depression, or related disorders during COVID-19 | Anxiety, depressive, or related disorder | DSM-V | 33.74 (12.64) | Female N=47 (67.1%) | White: 74% | Three-Item Loneliness Scale (TILS) | 12 weeks |
| Walker et al., 2020 | USA | RCT analysed as a prospective cohort study | 228 | Primary care patients with major depression | MDD | DSM-IV (SCID) | Low SCL: 48.6 (14.1) High SCL: 43.9 (12.4)<br><br>18-80 years | Low SCL N=149 Female: 73.2% | White: 80.5% (low SCL) 79.7% (high SCL) | Social Adjustment Scale (modified) | 1,3 and 6 months |

| | | | | | | | | | | | |
|---|---|---|---|---|---|---|---|---|---|---|---|
| | | | | | | | | High SCL N=79<br><br>Female: 77.2% | | | |
| Wielaard et al., 2017 | Netherlands | Longitudinal cohort study | 282 | Older adults with a depression diagnosis (NESDO) | MDD, dysthymia, minor depression | CIDI | 60+ years<br><br>70.6 (SD not provided) | NR | NR | 11-item loneliness scale | 2 years |

*NR: Not reported

**Table 2. Study findings of Near Misses**

| Author(s), Year | Setting | Main CMD outcome (s) | Main CMD outcome measure (s) | Adjusted for | Statistical Methods | Results of cross-sectional associations between loneliness and outcomes (Effect size, 95% CI, P-value) | Intervention (if applicable) | Conclusions |
|---|---|---|---|---|---|---|---|---|
| **A. General Population (care unknown) n=1** | | | | | | | | |
| Keller et al., 2023 | Medical Rehabilitation clinics | Depression | PHQ-2 | Age, gender, education, ICD-10 diagnosis | Serial mediation analysis using PROCESS macro | Loneliness and anxiety mediated the relationship between distress and depressive symptoms. Direct effect: standardized $\beta = 0.134$ ($p < 0.05$); indirect path through loneliness: $\beta = 0.176$ (95% CI [0.118–0.241]). | | Anxiety and loneliness as sustaining factors of depressive symptoms during the COVID-19 pandemic. |

| | | | | | | | | |
|---|---|---|---|---|---|---|---|---|
| **B. Using mental health services n=1** | | | | | | | | |
| Wielaard et al., 2017 | Mental health care institutes and general practitioners | Depression diagnosis | DSM-IV-TR criteria-based Composite International Diagnostic Interview (CIDI) | Baseline depression severity, neuroticism, age at depression onset, chronic diseases, loneliness | Mediation analysis, bootstrapping | Adjusted: Baseline loneliness significantly mediated the association between childhood abuse and depression diagnosis at T1 (B=0.18, 95% CI:0.04–0.38, $p < 0.05$). | | Loneliness alongside other factors explain a poor course of depression in older adults who reported childhood abuse. |
| **C. Receiving a specified clinical treatment (treatment response studies) n=4** | | | | | | | | |
| Jesus et al. 2024 | Community | Depression and anxiety symptoms | Geriatric Depression Scale-30 (GDS-30)<br><br>Geriatric Anxiety Inventory (GAI) | Age, sex, education level, marital status, living arrangement, medical conditions, baseline loneliness, baseline depressive symptoms (GDS-30), and baseline anxiety symptoms (GAI). | Primary Analysis: Linear mixed models (LMM) for repeated measures.<br><br>MEMORE: Mediation and Moderation analysis | Adjusted: Within the HEPPI (intervention) group, there was a significant indirect effect of social loneliness at T1 (3 months) on depressive symptoms reduction at T1 (3 months) (B = -1.52, SE = 0.19, 95% CI = -1.96, -1.22, $p < 0.001$). | Intervention: The Homebound Elderly People Psychotherapeutic Intervention (HEPPI) is a 10-week, home-based program for older adults with mild cognitive impairment and depressive/anxiety symptoms. It combines cognitive training, psychotherapeutic techniques (e.g., cognitive restructuring, | The intervention (HEPPI) led to lower depressive symptoms through a reduction in social loneliness. Participants in the HEPPI group showed a decrease in depressive symptoms because they felt less socially lonely after the intervention. |

| | | | | | | | | |
|---|---|---|---|---|---|---|---|---|
| | | | | | | | behavioral activation, mindfulness), and compensatory memory strategies | |
| O'Day et al. 2021 | Clinical referrals and community listings | Social anxiety | The Liebowitz Social Anxiety Scale-Self-Report (LSAS-SR) | Pre-treatment loneliness and its interaction with time | Multilevel linear models (MLMs) | <u>Adjusted:</u> Greater reduction in loneliness from pre-to post-treatment (T0 to T1), controlling for pre-treatment loneliness, were associated with lower levels of social anxiety on average during follow-up T2 (($\beta = -1.50$, $p < 0.001$). The interaction between time and loneliness change from pre- to post-treatment was not significant, $p > .05$. | Participants were receiving treatment for social anxiety disorder.<br><br>Cognitive-behavioural group therapy (CBGT)<br><br>Mindfulness-based stress reduction (MBSR) | Reductions in loneliness from pre- to post-treatment were significantly associated with lower levels of social anxiety on average during follow-up.<br>No significant interaction between time and social anxiety change. |
| Southward et al. 2022 | From Kentucky, USA | Depression, Anxiety | Overall Anxiety Severity and Impairment Scale (OASIS)<br>Overall Depression Severity and Impairment Scale (ODSIS) | Anxiety, depression, therapist effects | Hierarchical linear modelling (HLM) | Within-persons, higher loneliness was concurrently associated with higher anxiety ($r=0.22$, $p<0.01$) and depression ($r=0.31$, $p<0.01$) at each session. | The Unified Protocol for Transdiagnostic Treatment of Emotional Disorders (UP) is a modular, cognitive-behavioural therapy (CBT) approach designed to treat anxiety, depression, and related conditions. | |

| | | | | | | | | |
|---|---|---|---|---|---|---|---|---|
| Walker et al., 2020 | Primary care clinics | Depression severity | Hopkins Symptom Checklist (SCL-20) | Age, gender, neuroticism score, chronic disease score | Logistic regression, random effects models | <u>Adjusted:</u> Individuals who reported loneliness were 2.6 times more likely to fall into the higher depression severity group at baseline compared to those with lower loneliness scores (OR: 2.6, $p < 0.002$). | <u>Intervention:</u> Stepped Collaborative Care aiming to improve depression management in primary care settings by combining specialist support, patient education, and medication monitoring. | Loneliness emerged as one of the top psychosocial predictors, alongside comorbid panic disorder and childhood emotional abuse, for higher depression severity at baseline. |
